# To mask, or not to mask, Alice and Bob’s dating dilemma

**DOI:** 10.1101/2022.04.14.22273886

**Authors:** Zhanshan (Sam) Ma, Ya-Ping Zhang

## Abstract

Face masking in current COVID-19 pandemic seems to be a deceivingly simple decision-making problem due to its multifaceted nature. Questions arising from masking span biomedicine, epidemiology, physics, and human behaviors. While science has shown masks work generally, human behaviors (particularly under influences of politics) complicate the problem significantly given science generally assumes rationality and our minds are not always rational and/or honest. Minding minds, a legitimate concern, can also make masking legitimately confusing. To disentangle the potential confusions, particularly, the ramifications of irrationality and dishonesty, here we resort to evolutionary game theory. Specifically, we formulate and analyze the masking problem with a fictitious pair of young lovers, Alice and Bob, as a Sir Philip Sydney (SPS) evolutionary game, inspired by the handicap principle in evolutionary biology and cryptography figures in computer science. With the proposed ABD (Alice and Bob’s dating dilemma) as an asymmetric four-by-four strategic-form game, 16 strategic interactions were identified, and six of which may reach equilibriums with different characteristics such as separating, pooling, and polymorphic hybrid, being Nash, evolutionarily stable or neutrally stable. The six equilibrium types seem to mirror the diverse behaviors of mask believers, skeptics, converted, universal masking, voluntarily masking, coexisted and/or divided world of believers and skeptics. We suggest that the apparently simple ABD game is sufficiently general not only for studying masking policies for populations (*via* replicator dynamics), but also for investigating other complex decision-making problems with COVID-19 pandemic including lockdown *vs*. reopening, herd immunity *vs*. quarantines, and aggressive tracing *vs*. privacy protection.

## INTRODUCTION-Alice and Bob’s Dating Dilemma (ABD)

“*To Be, or Not to Be, That is the Question*” from Hamlet by William Shakespeare

The doubts and uncertainties surrounding masking *vs*. not-masking dilemma are worthy of careful examinations not only because the question *per se* is important but also it may act as a prototype for other potentially more challenging decision-making problems such as lockdowns *vs*. reopening, herd immunity *vs*. quarantines, and aggressive contact-tracing *vs*. privacy protection in facing the unprecedented challenges from the COVID-19 pandemic. Here we formulate and analyze the dilemma with a fictitious pair of young lovers, Alice and Bob, who were ‘professionally hackers’ before the COVID-19 pandemic (http://cryptocouple.com/).

After a few weeks of lockdown during the COVID-19 pandemic, Alice sent Bob a “miss you” message and they decided to meet somewhere. Alice could have two states: either healthy or infected by COVID-19. However, Bob had no clue on Alice’s state, and indeed, he did not even know his own health status. Before departing for the dating, Bob fell in a dilemma—should he wear a mask? He might be concerned with traditional social stigma such as being conceived as timid, unfavorable perception from Alice, and/or the infection risk. Alice might also fell in a dilemma, in particular, should she be honest to Bob about her health (COVID-19) status?

Time back to five centuries ago, British poet-soldier Sir Philip Sidney (1554-1586), when fatally injured in a battle, with the immortal words “*thy necessity is greater than mine*”, passed his water bottle to one fellow soldier who was also a casualty. Whether or not this well-known English story is true is less relevant for the topic of this article since the other scenarios are well covered by famous evolutionary biologist John Maynard-Smith (1920-2004), who transformed the story into a rather successful evolutionary game for investigating the honesty in animal communications. With his words, “*the story deserves to be true, although it was based on the claims of a close friend of Sir Philip Sidney*”. Maynard-Smith (1991, 2003) formulated the Sir Philip Sidney (SPS) game with an objective to resolve a then hotly debated hypothesis—the *handicap principle* first proposed by Zahavi (1975, 1997), who tackled a fundamental problem in evolutionary biology with deep humanity implications. The handicap principle maintains that animal signaling (communication) must be costly to be reliable in the existence of conflicts of interests. For example, the tail of a male peacock is costly because it attracts the attention of its predator, but it also acts as a reliable (honest) signal to female to demonstrate the male’s fitness (genetic quality). Similarly, in human society, the luxury goods must be costly to demonstrate its value. The handicap principle also shed light on a question of humanity: are we human beings the only creatures that can lie? If not, how the honest signaling was evolved in animal world?

The Sir Philip Sidney (SPS) game is an action-response game and proceeds in two stages with two participants (Huttegger & Zollman 2010, Whitmeyer 2020): signaler (the message sender or Sir Philip Sidney’s comrade in this case) may send a request to donor (responder or message receiver: Sir Philip) for water bottle and the responder may or may not respond by donating the bottle. Signaler could be in one of two states: healthy with probability of (1–*m*), wounded (needy) (*m*). If he receives the water (resource) from the responder, he will survive. Otherwise, his survival probability is (1–*b*) if healthy, and (1–*a*) if wounded (*a*>*b*). In addition, the signaler may need to pay for the cost (*c*) of sending the request. On the responder side, he can either donate the water or do nothing. In the former case, his survival probability is (1–*d*), and in the latter case, the survival probability is unchanged. Note that the ‘cost’ refers to ‘*strategic cost*’ in general, which may include the consequence from dishonest signaling, such as the increased risk of infection or decreased survival probability in the case of ABD problem.

There is a relatedness parameter (*k*), which is defined as the proportion of shared ‘genes’ between a signaler and a potential responder. Both the sender and responder behave to maximize their own ‘inclusive fitness’ (from *kinship* theory), which are the *k* times the payoff of the other player plus his or her own payoff. This parameter is critical for the SPS game because it defines the level of common interests (the opposite of conflicts of interests) and therefore potentially has a critical impact on the reliability of signaling (Maynard Smith & Harper 2003, Cooper *et al* 2018). When the *k*-value is larger, the players are more closely related, the signaler is more likely to signal honestly and the donor is more likely to donate. In our ABD model, the relatedness parameter (*k*) represents the closeness of the love relationship between Alice and Bob, and measures their shared interests in masking-or-not decision-making.

The SPS game was initially formulated to demonstrate the equilibriums underlying the handicap principle, *i.e*., signaling must be costly to be honest in the existence of conflict of interests. The equilibriums (*see* Box 3) could be evolutionarily stable, neural or unstable. For example, if the signaler only signals when in need and if the responder only donates the water in response to a signal, there is a signaling Nash equilibrium. It could be easily derived that *c=*(*b–rd*) is the minimum cost to maintain this equilibrium, *i.e*., the honest signaling. When *b*<*rd*, cost-free signaling is possible (*c*≤0); then, the signaler may signal (lie) even if he is not in need (Maynard Smith & Harper 2003). It was rumored that Maynard Smith (1991) designed the simple SPS game when he was frustrated with the overly complexity of Grafen’s (1990) game model for sexual selection, which for the first time demonstrated the validity of Zahavi’s (1975, 1991) handicap principle, but the mathematics was slightly too complex. The SPS is indeed simpler than Grafen’s model, still demonstrated the theoretical feasibility of the handicap principle. Nevertheless, in the consequent two decades, researchers quickly discovered some initially ignored subtleties of SPS game, and the expanded studies on those intricacies not only significantly advanced the field of animal communications (signaling), but also made the SPS game one of the most important game theoretic models in evolutionary biology. Whereas the well-known prisoner’s dilemma has been a *de* f*acto* standard model for investigating the *cooperation* in evolutionary biology, it is the SPS game that acts as a *de facto* standard for studying the *communication*.

From a broad perspective, it was argued that the three processes, *i.e*., competition, cooperation and communication, are three fundamental manifestations of animal behaviors under the natural selection that drives the evolution of organisms (Ma 2015a, 2015b). While the roles of competition have been extensively investigated ever since Darwin’s evolutionary theory, those of cooperation have also been widely approached since the 1960s thanks to pioneering works such as Hamilton (1964). The study of cooperation took off in the 1980s and it also popularized the applications of prisoner’s dilemma (PD) game (Axelrod & Hamilton 1980). Zahavi’s handicap principle (1975, 1997) suggested that communication was a missing piece in Darwin’s evolutionary theory, but it was initially rejected until Grafen (1990) and Maynard-Smith (1991, 2003) game-theoretic analyses in the early 1990s formally demonstrated its theoretical validity. Both PD and SPS games belong to the so-termed evolutionary games, which can be considered as a marriage between Darwin’s evolutionary theory and game theory, and in fact, it was George Price and Maynard Smith (1973) who invented the evolutionary game theory (EGT), while they were inspired by the observations of animal communication behavior. Today, the EGT has become one of the most actively pursued branches of game theory and has found applications well beyond its original field of evolutionary biology. In addition, the communication behaviors are not limited to animals and microbes, and indeed, plants communicate too (Baluska *et al*. 2006). Obviously, we humans are likely to be the most capable creatures on the earth planet, in both communicating honestly and deceivably. Today, in facing the COVID-19 pandemic, being one of the biggest challenges recent decades, we believe it is worthy to carefully examine the implications of the dilemma faced by Alice and Bob because the problem is sufficiently general for us to gain insights into other more severe challenges posed by the pandemic, besides the theoretical and practical implications of the dilemma *per se*.

The objective of this study is to present a game-theoretic approach for investigating complex decision-making problems in devising public health policies, particularly in dealing with the COVID-19 pandemic, by demonstrating its application to solve the problem of Alice-Bob dating dilemma (ABD) outlined previously. Specifically, we transform the ABD as a SPS evolutionary game. Furthermore, we adopt arguably the most general form of the SPS game, as expanded by Bergstrom & Lachmann (1997, 1998), Huttegger & Zollman (2010) and Whitmeyer (2020). These existing works significantly expanded the domain of SPS game by considering all possible strategies between the players. In particular, the results of *pooling equilibriums* (in which signalers in different states “pool” and send the same message, or no communications between players in this article) or *strategic inattention* can be particularly powerful for analyzing our ABD and other COVID-19 problems because of the need for dealing with privacy concerns. An even more compelling reason is that Huttegger & Zollman (2010) analyses with *replicator dynamics* actually puts the SPS game onto population scale. In other words, it can be used to study the masking policies for a region or country’s whole population, rather than for Alice and Bob only. In addition, the *polymorphism*, where players mix between being honest and being deceptive while the signaling costs can be very low, can also be more realistic in capturing the enormous controversies and uncertainties surrounding mask wearing today. To the best of our knowledge, this article should be the first application of game theory for investing masking strategies. Furthermore, our approach is particularly suitable for dealing with the issues of irrationality and deception (dishonesty) in modeling masking strategies because (*i*) evolutionary game theory is not plagued by the *rationality* assumption as in classic game theory and (*ii*) the handicap principle and its evolutionary game model (SPS game) is designed to effectively deal with possible deception (dishonesty) in communications.

## Material and Methods

This game-theoretic study does not involve any experimental/survey experiments, except for simulations. For this, a standard section of material and methods is hardly applicable for organizing this article. In the **Introduction** section, the problem of masking strategy is introduced with a fictitious pair of young lovers (Alice and Bob) as Alice and Bob’s dating Dilemma (ABD), and the problem is formulated as an extended Sir Philip Sydney (SPS) game, originally invented in evolutionary biology. In the consequent **Results** section, the analytic and simulation analyses of the ABD are presented, respectively. The article is completed with the section of **Conclusions, Discussion and Perspective**.

## RESULTS—Analysis and Simulation of ABD

### Formulation of the ABD game

Table 1 illustrates the formulation of ABD (Alice and Bob’s dating dilemma) problem as a SPS game. It listed the four possible options (strategies) Alice and Bob each may have and their 16 possible interactions, mirrored the options of Sir Philip and his comrade in the traditional SPS game. Fig 1 further displays the ABD as a decision-tree and strategy matrix. In consideration of the counter-intuitive nature of the strategy set, Box 1 further exposes the formulation and transformation of the ABD game from the classic SPS game, including the three elements (*who, what*, and *how*) of the SPS/ABD game. Box 2 exposes the parameters of the SPS/ABD game.

**Table 1.** From Sir Philip Sidney (SPS) game to Alice-Bob Dating Dilemma (ABD) game*

| Sir Philip Sydney (SPS) Game and its Extensions<br>(Maynard-Smith 1992, Huttegger & Zollman 2010, Ma & Krings 2011, Whitmeyer 2020) |  |  |  |  |
| --- | --- | --- | --- | --- |
| Sender (Signaler) Strategies |  | Responder (Receiver or Donor) Strategies |  |  |
| S<sub>1</sub>: Signal only if healthy |  | R<sub>1</sub>: Donate only if no signal |  |  |
| S<sub>2</sub>: Signal only if needy |  | R<sub>2</sub>: Donate only if signal |  |  |
| S<sub>3</sub>: Never signal |  | R<sub>3</sub>: Never donate |  |  |
| S<sub>4</sub>: Always signal |  | R<sub>4</sub>: Always donate |  |  |
| Alice and Bob’s Dating Dilemma (ABD) Formulated as an Extended SPS Game |  |  |  |  |
| Alice’s Strategies |  | Bob’s Strategies |  |  |
| A<sub>1</sub>: Not masking only if healthy |  | B<sub>1</sub>: Not masking only if Alice masks |  |  |
| A<sub>2</sub>: Not masking only if infected |  | B<sub>2</sub>: Not masking only if Alice does not mask |  |  |
| A<sub>3</sub>: Always masking |  | B<sub>3</sub>: Always masking |  |  |
| A<sub>4</sub>: Never masking |  | B<sub>4</sub>: Never masking |  |  |
| For examples, A<sub>1</sub>: Alice’s strategies:<br>Not masking = Signaling: “I am healthy” (Not infected) with probability (1-*m*).<br>Masking = Not Signaling: “I am infected with COVID-19” (needy) with probability *m*. |  | For examples, B<sub>1</sub>: Bob’s strategies:<br>Not Masking=Responding to Alice’s signal (not masking) with potential loss of *d* (get infected).<br>Masking=Not responding to Alice’s signal (of not masking) without loss. |  |  |
| The 4x4 combinatorial asymmetric strategies of the discrete extended SPS/ABD game with the letters (A-P) representing for possible strategy pairs as shown in **Fig 1** |  |  |  |  |
| Responder’s strategies<br>(Bob’s strategies)<br>Sender strategies<br>(Alice’s Strategies) | R<sub>1</sub> (B<sub>1</sub>)<br>Donate only if<br>no signal<br>(Not masking<br>only if Alice<br>masks) | R<sub>2</sub> (B<sub>2</sub>)<br>Donate only if<br>signal<br>(Not masking only<br>if Alice does not<br>mask) | R<sub>3</sub> (B<sub>3</sub>)<br>Never<br>Donate<br>(Always<br>masking) | R<sub>4</sub> (B<sub>4</sub>)<br>Always<br>donate<br>(Never<br>masking) |
| S<sub>1</sub> (A<sub>1</sub>): Signal only if healthy<br>(Not masking only if healthy) | A | B | C | D |
| S<sub>2</sub> (A<sub>2</sub>): Signal only if needy<br>(Not masking only if infected) | E | F | G | H |
| S<sub>3</sub> (A<sub>3</sub>): Never signal<br>(Always masking) | I | J | K | L |
| S<sub>4</sub> (A<sub>4</sub>): Always signal<br>(Never masking) | M | N | O | P |

\*Notes: See Box 1, and Fig 1 for the exposition of the ABD game formulation/transformation from the extended SPS, and Box 2 for the interpretations of the SPS/ABD game parameters.

**Fig 1.**
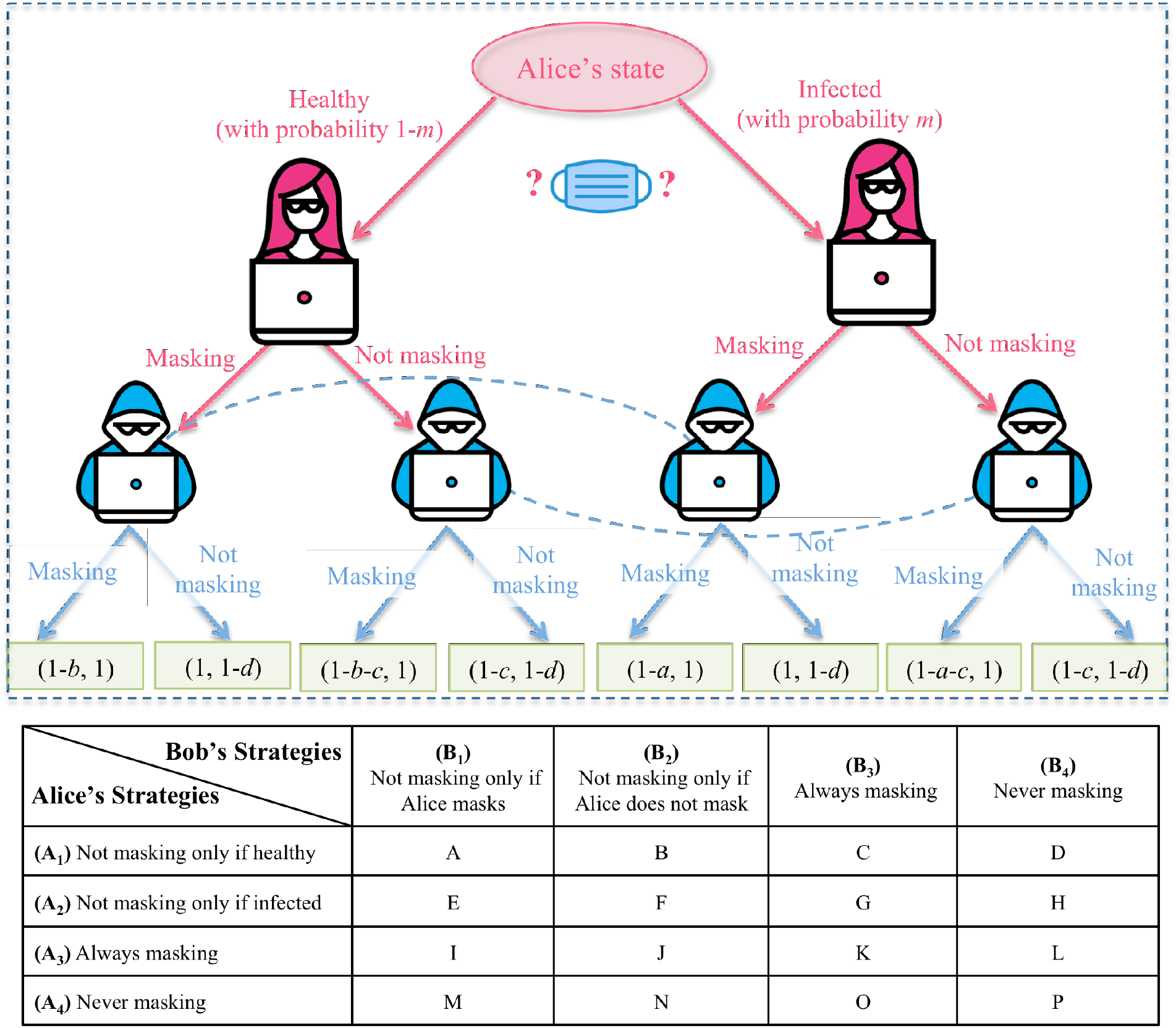
A decision-tree of ABD (Alice-Bob masking dilemma) formulated as a 4-by-4 asymmetrical SPS (Sir Philip Sidney) game: The terminal nodes show the payoffs of Alice and Bob, respectively. The dotted lines depict the Alice’s information sets, but Bob may not be able to distinguish the decision nodes connected by the dotted lines. In other words, the dotted lines indicate that Bob does not know Alice’s health status ahead of their dating.

#### Box 1.

Three Key Elements (*Who, What* and *How*) to Formulate SPS/ABD Games and the Exposition of the Transformation from SPS to ABD game

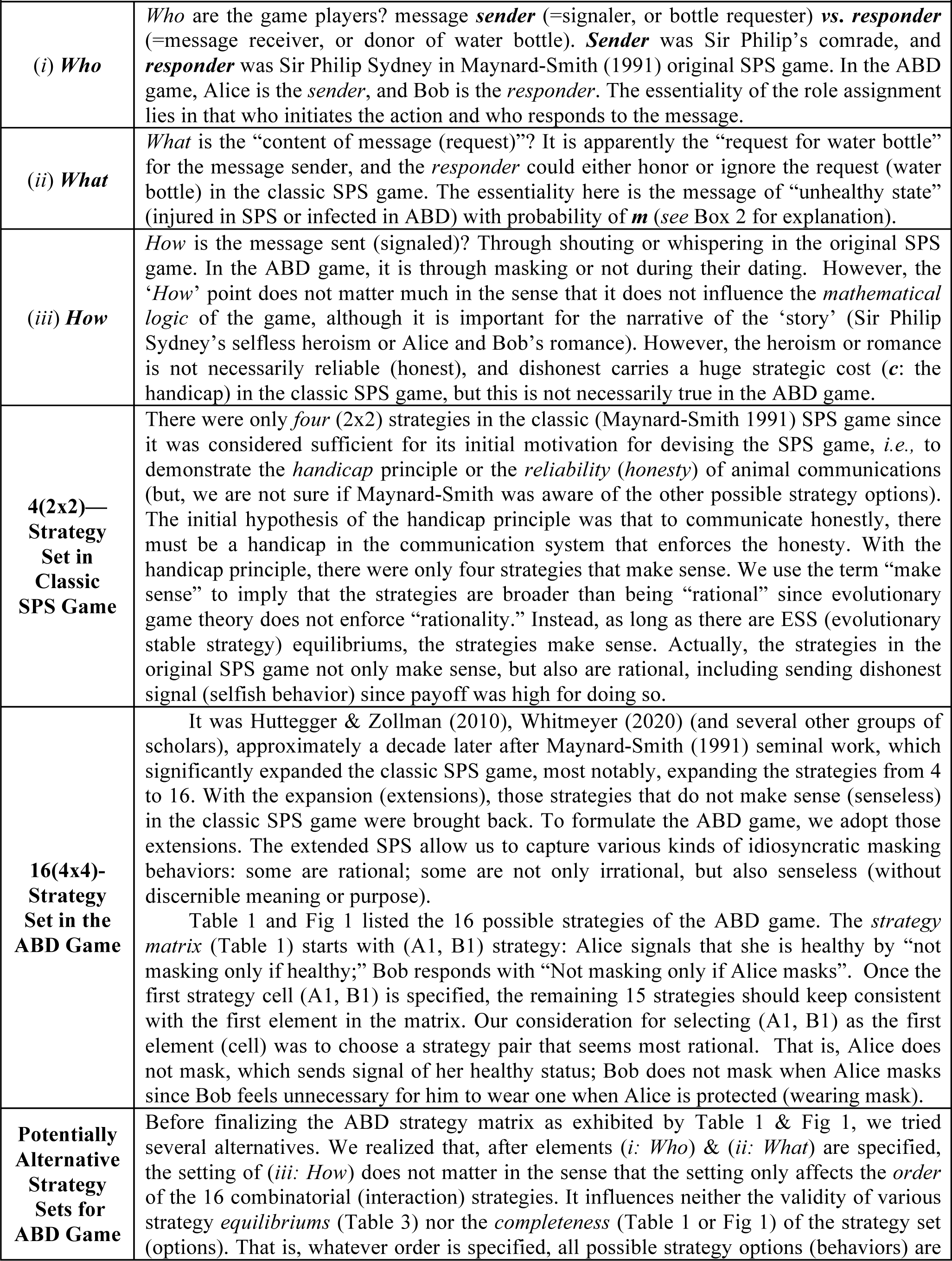
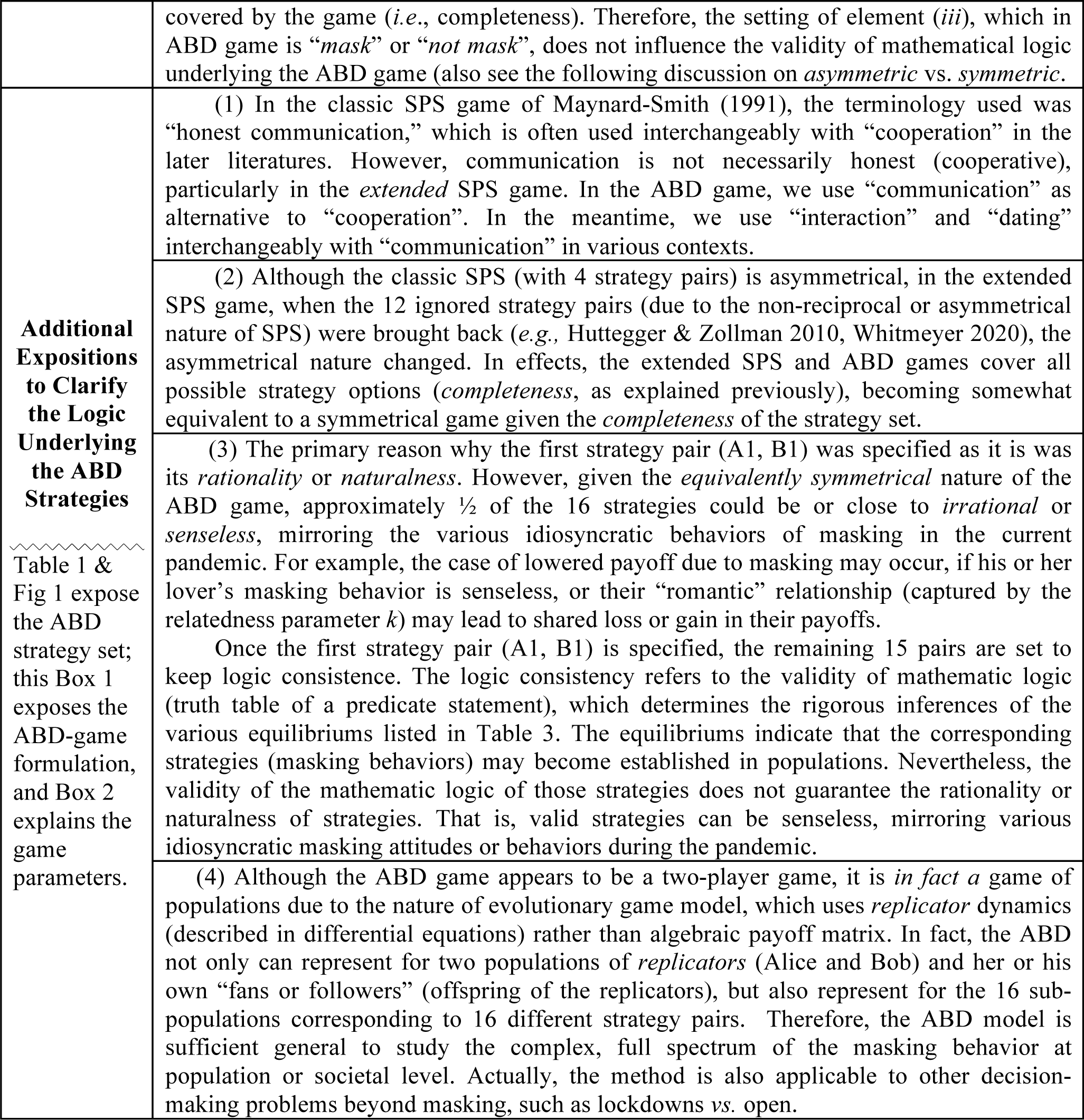

| **Box 1. Three Key Elements (*Who*, *What* and *How*) to Formulate SPS/ABD Games and the Exposition of the Transformation from SPS to ABD game** |  |
| --- | --- |
| **(i) *Who*** | *Who* are the game players? message **sender** (=signaler, or bottle requester) **vs.** **responder** (=message receiver, or donor of water bottle). **Sender** was Sir Philip's comrade, and **responder** was Sir Philip Sydney in Maynard-Smith (1991) original SPS game. In the ABD game, Alice is the *sender*, and Bob is the *responder*. The essentiality of the role assignment lies in that who initiates the action and who responds to the message. |
| **(ii) *What*** | *What* is the "content of message (request)"? It is apparently the "request for water bottle" for the message sender, and the *responder* could either honor or ignore the request (water bottle) in the classic SPS game. The essentiality here is the message of "unhealthy state" (injured in SPS or infected in ABD) with probability of *m* (see Box 2 for explanation). |
| **(iii) *How*** | *How* is the message sent (signaled)? Through shouting or whispering in the original SPS game. In the ABD game, it is through masking or not during their dating. However, the '*How*' point does not matter much in the sense that it does not influence the *mathematical logic* of the game, although it is important for the narrative of the 'story' (Sir Philip Sydney's selfless heroism or Alice and Bob's romance). However, the heroism or romance is not necessarily reliable (honest), and dishonest carries a huge strategic cost (*c*: the handicap) in the classic SPS game, but this is not necessarily true in the ABD game. |
| **4(2x2)—**<br>**Strategy**<br>**Set in**<br>**Classic**<br>**SPS Game** | There were only *four* (2x2) strategies in the classic (Maynard-Smith 1991) SPS game since it was considered sufficient for its initial motivation for devising the SPS game, *i.e.*, to demonstrate the *handicap* principle or the *reliability* (*honesty*) of animal communications (but, we are not sure if Maynard-Smith was aware of the other possible strategy options). The initial hypothesis of the handicap principle was that to communicate honestly, there must be a handicap in the communication system that enforces the honesty. With the handicap principle, there were only four strategies that make sense. We use the term "make sense" to imply that the strategies are broader than being "rational" since evolutionary game theory does not enforce "rationality." Instead, as long as there are ESS (evolutionary stable strategy) equilibriums, the strategies make sense. Actually, the strategies in the original SPS game not only make sense, but also are rational, including sending dishonest signal (selfish behavior) since payoff was high for doing so. |
| **16(4x4)-**<br>**Strategy**<br>**Set in the**<br>**ABD Game** | It was Huttegger & Zollman (2010), Whitmeyer (2020) (and several other groups of scholars), approximately a decade later after Maynard-Smith (1991) seminal work, which significantly expanded the classic SPS game, most notably, expanding the strategies from 4 to 16. With the expansion (extensions), those strategies that do not make sense (senseless) in the classic SPS game were brought back. To formulate the ABD game, we adopt those extensions. The extended SPS allow us to capture various kinds of idiosyncratic masking behaviors: some are rational; some are not only irrational, but also senseless (without discernible meaning or purpose).<br>Table 1 and Fig 1 listed the 16 possible strategies of the ABD game. The *strategy matrix* (Table 1) starts with (A1, B1) strategy: Alice signals that she is healthy by "not masking only if healthy;" Bob responds with "Not masking only if Alice masks". Once the first strategy cell (A1, B1) is specified, the remaining 15 strategies should keep consistent with the first element in the matrix. Our consideration for selecting (A1, B1) as the first element (cell) was to choose a strategy pair that seems most rational. That is, Alice does not mask, which sends signal of her healthy status; Bob does not mask when Alice masks since Bob feels unnecessary for him to wear one when Alice is protected (wearing mask). |
| **Potentially**<br>**Alternative**<br>**Strategy**<br>**Sets for**<br>**ABD Game** | Before finalizing the ABD strategy matrix as exhibited by Table 1 & Fig 1, we tried several alternatives. We realized that, after elements (*i: Who*) & (*ii: What*) are specified, the setting of (*iii: How*) does not matter in the sense that the setting only affects the *order* of the 16 combinatorial (interaction) strategies. It influences neither the validity of various strategy *equilibriums* (Table 3) nor the *completeness* (Table 1 or Fig 1) of the strategy set (options). That is, whatever order is specified, all possible strategy options (behaviors) are |
|  | covered by the game (*i.e.*, completeness). Therefore, the setting of element (*iii*), which in ABD game is “*mask*” or “*not mask*”, does not influence the validity of mathematical logic underlying the ABD game (also see the following discussion on *asymmetric* vs. *symmetric*). |
| **Additional Expositions to Clarify the Logic Underlying the ABD Strategies**<br>~~~~~<br>Table 1 & Fig 1 expose the ABD strategy set; this Box 1 exposes the ABD-game formulation, and Box 2 explains the game parameters. | (1) In the classic SPS game of Maynard-Smith (1991), the terminology used was “honest communication,” which is often used interchangeably with “cooperation” in the later literatures. However, communication is not necessarily honest (cooperative), particularly in the *extended* SPS game. In the ABD game, we use “communication” as alternative to “cooperation”. In the meantime, we use “interaction” and “dating” interchangeably with “communication” in various contexts. |
|  | (2) Although the classic SPS (with 4 strategy pairs) is asymmetrical, in the extended SPS game, when the 12 ignored strategy pairs (due to the non-reciprocal or asymmetrical nature of SPS) were brought back (*e.g.*, Huttegger & Zollman 2010, Whitmeyer 2020), the asymmetrical nature changed. In effects, the extended SPS and ABD games cover all possible strategy options (*completeness*, as explained previously), becoming somewhat equivalent to a symmetrical game given the *completeness* of the strategy set. |
|  | (3) The primary reason why the first strategy pair (A1, B1) was specified as it is was its *rationality* or *naturalness*. However, given the *equivalently symmetrical* nature of the ABD game, approximately $\frac{1}{2}$ of the 16 strategies could be or close to *irrational* or *senseless*, mirroring the various idiosyncratic behaviors of masking in the current pandemic. For example, the case of lowered payoff due to masking may occur, if his or her lover’s masking behavior is senseless, or their “romantic” relationship (captured by the relatedness parameter *k*) may lead to shared loss or gain in their payoffs.<br>Once the first strategy pair (A1, B1) is specified, the remaining 15 pairs are set to keep logic consistence. The logic consistency refers to the validity of mathematic logic (truth table of a predicate statement), which determines the rigorous inferences of the various equilibriums listed in Table 3. The equilibriums indicate that the corresponding strategies (masking behaviors) may become established in populations. Nevertheless, the validity of the mathematic logic of those strategies does not guarantee the rationality or naturalness of strategies. That is, valid strategies can be senseless, mirroring various idiosyncratic masking attitudes or behaviors during the pandemic. |
|  | (4) Although the ABD game appears to be a two-player game, it is *in fact* a game of populations due to the nature of evolutionary game model, which uses *replicator* dynamics (described in differential equations) rather than algebraic payoff matrix. In fact, the ABD not only can represent for two populations of *replicators* (Alice and Bob) and her or his own “fans or followers” (offspring of the replicators), but also represent for the 16 sub-populations corresponding to 16 different strategy pairs. Therefore, the ABD model is sufficient general to study the complex, full spectrum of the masking behavior at population or societal level. Actually, the method is also applicable to other decision-making problems beyond masking, such as lockdowns vs. open. |

#### Box 2.

Interpretations of the parameters (*a, b, c, d, m* & *k*) in the SPS/ABD Games

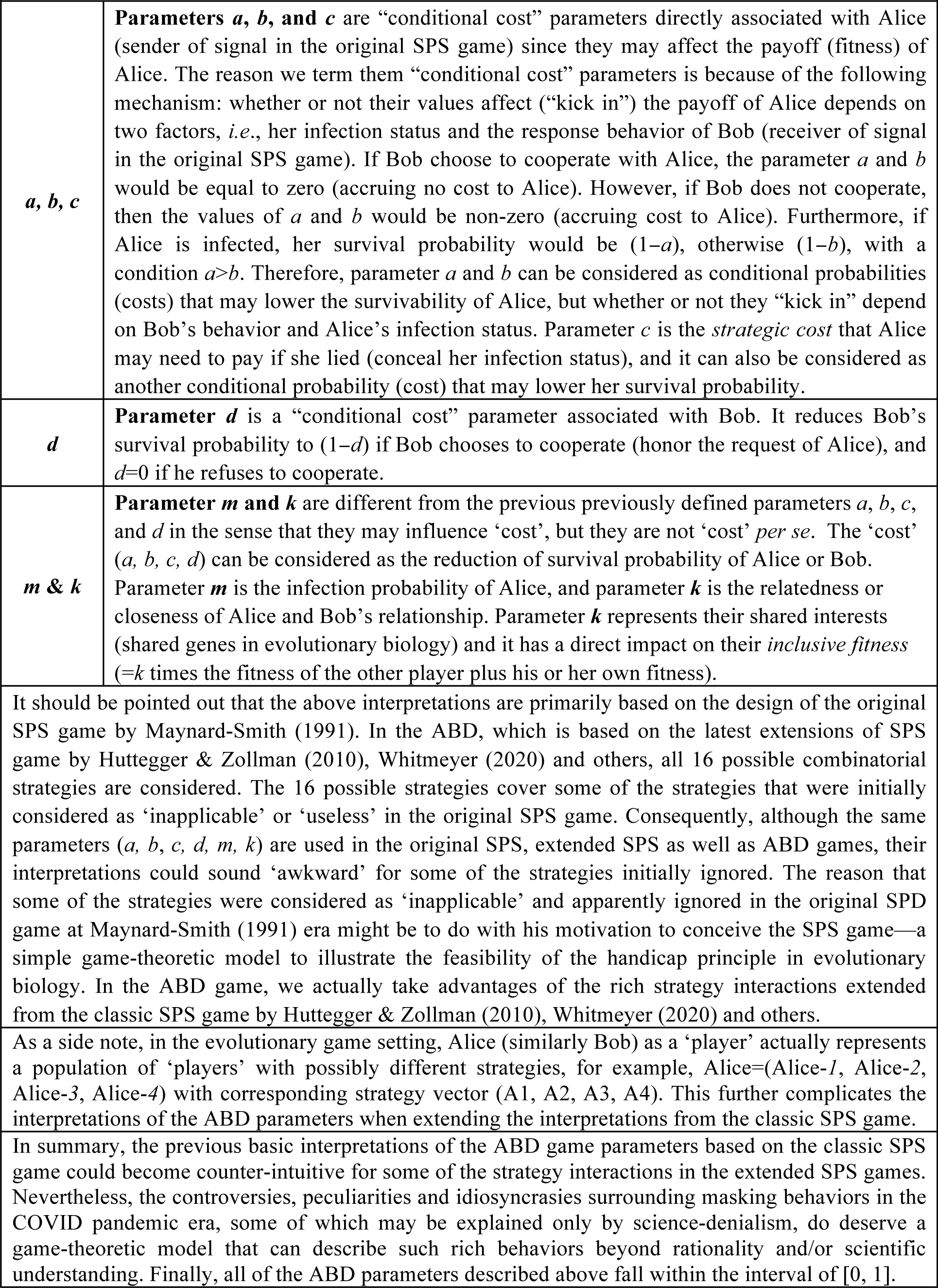

| **Box 2. Interpretations of the parameters ($a, b, c, d, m$ & $k$) in the SPS/ABD Games** |  |
| --- | --- |
| **$a, b, c$** | **Parameters $a, b$, and $c$** are “conditional cost” parameters directly associated with Alice (sender of signal in the original SPS game) since they may affect the payoff (fitness) of Alice. The reason we term them “conditional cost” parameters is because of the following mechanism: whether or not their values affect (“kick in”) the payoff of Alice depends on two factors, *i.e.*, her infection status and the response behavior of Bob (receiver of signal in the original SPS game). If Bob choose to cooperate with Alice, the parameter $a$ and $b$ would be equal to zero (accruing no cost to Alice). However, if Bob does not cooperate, then the values of $a$ and $b$ would be non-zero (accruing cost to Alice). Furthermore, if Alice is infected, her survival probability would be $(1-a)$, otherwise $(1-b)$, with a condition $a > b$. Therefore, parameter $a$ and $b$ can be considered as conditional probabilities (costs) that may lower the survivability of Alice, but whether or not they “kick in” depend on Bob’s behavior and Alice’s infection status. Parameter $c$ is the *strategic cost* that Alice may need to pay if she lied (conceal her infection status), and it can also be considered as another conditional probability (cost) that may lower her survival probability. |
| **$d$** | **Parameter $d$** is a “conditional cost” parameter associated with Bob. It reduces Bob’s survival probability to $(1-d)$ if Bob chooses to cooperate (honor the request of Alice), and $d=0$ if he refuses to cooperate. |
| **$m$ & $k$** | **Parameter $m$ and $k$** are different from the previous previously defined parameters $a, b, c$, and $d$ in the sense that they may influence ‘cost’, but they are not ‘cost’ *per se*. The ‘cost’ ($a, b, c, d$) can be considered as the reduction of survival probability of Alice or Bob. Parameter $m$ is the infection probability of Alice, and parameter $k$ is the relatedness or closeness of Alice and Bob’s relationship. Parameter $k$ represents their shared interests (shared genes in evolutionary biology) and it has a direct impact on their *inclusive fitness* ($=k$ times the fitness of the other player plus his or her own fitness). |
| It should be pointed out that the above interpretations are primarily based on the design of the original SPS game by Maynard-Smith (1991). In the ABD, which is based on the latest extensions of SPS game by Huttegger & Zollman (2010), Whitmeyer (2020) and others, all 16 possible combinatorial strategies are considered. The 16 possible strategies cover some of the strategies that were initially considered as ‘inapplicable’ or ‘useless’ in the original SPS game. Consequently, although the same parameters ($a, b, c, d, m, k$) are used in the original SPS, extended SPS as well as ABD games, their interpretations could sound ‘awkward’ for some of the strategies initially ignored. The reason that some of the strategies were considered as ‘inapplicable’ and apparently ignored in the original SPD game at Maynard-Smith (1991) era might be to do with his motivation to conceive the SPS game—a simple game-theoretic model to illustrate the feasibility of the handicap principle in evolutionary biology. In the ABD game, we actually take advantages of the rich strategy interactions extended from the classic SPS game by Huttegger & Zollman (2010), Whitmeyer (2020) and others. | |
| As a side note, in the evolutionary game setting, Alice (similarly Bob) as a ‘player’ actually represents a population of ‘players’ with possibly different strategies, for example, Alice=(Alice-1, Alice-2, Alice-3, Alice-4) with corresponding strategy vector (A1, A2, A3, A4). This further complicates the interpretations of the ABD parameters when extending the interpretations from the classic SPS game. |  |
| In summary, the previous basic interpretations of the ABD game parameters based on the classic SPS game could become counter-intuitive for some of the strategy interactions in the extended SPS games. Nevertheless, the controversies, peculiarities and idiosyncrasies surrounding masking behaviors in the COVID pandemic era, some of which may be explained only by science-denialism, do deserve a game-theoretic model that can describe such rich behaviors beyond rationality and/or scientific understanding. Finally, all of the ABD parameters described above fall within the interval of $[0, 1]$. | |

In Table 1 the top section is the Sir Philip Sidney (SPS) game proposed by Maynard-Smith (1991) and further extended by Huttegger & Zollman (2010) and Whitmeyer (2020). In formulating the ABD as a SPS game, we adopted slightly different symbols with the original SPS game, with Alice for ‘Signaler’ (‘Sender’) and Bob for ‘Responder’ (‘Donor’). As explained in Table 1 and Fig 1, we map ‘not masking’ in the ABD to ‘signaling’ in the original SPS game transmitting signal—”I am healthy or not infected with COVID-19.” This is equivalent to use ‘masking’ for transmitting signal—I am infected with COVID-19. Regarding Bob’s response, ‘not masking only if Alice masks’ is translated into ‘donate only if no signal,’ and ‘not masking only if Alice does not mask’ is translated into ‘donate only if signal.’

Obviously, since all 16 strategies from the 4-by-4 combinatorial options of Alice and Bob’s interactions are covered by the mapping to a SPS game, specific mapping between the strategies of ABD and existing SPS game is less important in analyzing the games. Nothing is really special about the mapping scheme we adopt in Table 1 other than considering/accommodating intuitions. For example, the very first mapping ‘not masking’ in the ABD game to ‘signaling’ in SPS game was in consideration of an intuition (opinion) of some people, *i.e*., there is not a need to mask if I am healthy, transmitting a signal of the healthy state. However, others may not agree with this opinion, and the alterative opinions (intuitions) are covered by other strategies in our translation scheme illustrated in Table 1 and Fig 1.

The bottom sections of Table 1 and Fig 1 assign a series of labels (A-P) for 16 combinatorial strategies of Alice and Bob, which makes it convenient for discussing the analytical results of the ABD game such as its equilibriums (Table 2) and payoff (fitness) (Table 3). Box 3 and Box 4 introduce some essential terminologies used for discussing the equilibriums and payoff (fitness) of the ABD game. *See* Box 1 for further clarifications of the remaining issues regarding the formulation of the ABD game, as well as the exposition of the ABD parameters (Box 2).

**Table 2.** The necessary conditions and stabilities of various equilibriums with the ABD game

| **Equilibrium Type**<br>(see Box 3) | **Necessary conditions for the**<br>**existence of the equilibriums**<br>(see Box 2 for the ABD parameters) | **Stability**<br>**Property**<br>(see Box 4) |
| --- | --- | --- |
| **F**—Signaling ESS (Separating equilibrium) | $a > c + kd > b$ and $a > \frac{d}{k} > b$ | ESS (Evolutionary Stable Strategy) and is Asymptotically stable. |
| **L**—Pooling equilibrium | $d < k[ma + (1 - m)b]$ | Neutrally stable |
| **A**—Nash equilibrium (Apparently rational equilibrium) | $a > kd - c > b$ and $a > \frac{d}{k} > b$ | Asymptotically stable |
| **\*J and K** are behaviorally equivalent.<br>$(A_3, (1 - \lambda)B_2 + \lambda B_3)$<br>—Pooling equilibrium | $d > k[ma + (1 - m)b]$<br>and $\lambda \geq 1 - \frac{c}{a - kd}$ | Neutrally stable |
| **\*I and L** are behaviorally equivalent.<br>$(A_3, (1 - \mu)B_1 + \mu B_4)$<br>—Pooling equilibrium | $d < k[ma + (1 - m)b]$<br>and $\mu \geq 1 - \frac{c}{kd - b}$ | Neutrally stable |
| $(\lambda A_2 + (1 - \lambda)A_4, \mu B_2 + (1 - \mu)B_3)$<br>—Polymorphic Hybrid equilibrium<br>$\lambda = \frac{k[ma + (1 - m)b] - d}{(1 - m)(kb - d)}$ &<br>$\mu = \frac{c}{b - kd}$ | $a > \frac{d}{k} > b$ & $b - kd > c$ &<br>$d > k[ma + (1 - m)b]$ | Lyapunov stable<br>(Asymptotically stable or unstable) |

*\*J & K* mean that $(A_3, B_2)$ and $(A_3, B_3)$ are behaviorally equivalent. Similarly, *I & L* mean that $(A_3, B_1)$ and $(A_3, B_4)$ are behaviorally equivalent. See Box 3 for the interpretations.

**Table 3.** The expected payoffs for Alice and Bob at the equilibriums of the ABD game

| Equilibrium Type (Box 3) | Alice's Payoff (Fitness) | Bob's Payoff (Fitness) |
| --- | --- | --- |
| **A** — (A<sub>1</sub>: Not masking only if healthy; B<sub>1</sub>: Not masking only if Alice masks) | $1 - b - c + k + m(b + c - dk)$ | $1 + k(1 - b - c) + m(kb + kc - d)$ |
| **B** — (A<sub>1</sub>: Not masking only if healthy; B<sub>2</sub>: Not masking only if Alice does not mask) | $1 - c + k(1 - d) + m(-a + c + kd)$ | $1 + k(1 - c) - d + m(-ak + ck + d)$ |
| **C** — (A<sub>1</sub>: Not masking only if healthy; B<sub>3</sub>: Always masking) | $1 - b - c + k + m(-a + b + c)$ | $1 + k(1 - b - c) + mk(-a + b + c)$ |
| **D** — (A<sub>1</sub>: Not masking only if healthy; B<sub>4</sub>: Never masking) | $1 - c + k(1 - d) + mc$ | $1 - d + k(1 - c) + mkc$ |
| **E** — (A<sub>2</sub>: Not masking only if infected; B<sub>1</sub>: Not masking only if Alice masks) | $1 + k(1 - d) + m(-a - c + kd)$ | $1 - d + k + m(-ka - kc + d)$ |
| **F** — (A<sub>2</sub>: Not masking only if infected; B<sub>2</sub>: Not masking only if Alice does not mask) | $1 - b + k + m(b - c - kd)$ | $1 - bk + k + m(bk - ck - d)$ |
| **G** — (A<sub>2</sub>: Not masking only if infected; B<sub>3</sub>: Always masking) | $1 - b + k + m(-a + b - c)$ | $1 + k(1 - b) + mk(-a + b - c)$ |
| **H** — (A<sub>2</sub>: Not masking only if infected; B<sub>4</sub>: Never masking) | $1 + k(1 - d) - mc$ | $1 - d + k - mkc$ |
| **I** — (A<sub>3</sub>: Always masking; B<sub>1</sub>: Not masking only if Alice masks) | $1 + k(1 - d)$ | $1 - d + k$ |
| **L** — (A<sub>3</sub>: Always masking; B<sub>4</sub>: Never masking) |  |  |
| **J** — (A<sub>3</sub>: Always masking; B<sub>2</sub>: Not masking only if Alice does not mask) | $1 - b + k + m(-a + b)$ | $1 + k(1 - b) + mk(-a + b)$ |
| **K** — (A<sub>3</sub>: Always masking; B<sub>3</sub>: Always masking) |  |  |
| **M** — (A<sub>4</sub>: Always masking; B<sub>1</sub>: Not masking only if Alice masks) | $1 - b - c + k + m(-a + b)$ | $1 + k(1 - b - c) + mk(-a + b)$ |
| **O** — (A<sub>4</sub>: Always masking; B<sub>3</sub>: Always masking) |  |  |
| **N** — (A<sub>4</sub>: Always masking; B<sub>2</sub>: Not masking only if Alice does not mask) | $1 - c + k(1 - d)$ | $1 - d + k(1 - c)$ |
| **P** — (A<sub>4</sub>: Always masking; B<sub>4</sub>: Never masking) |  |  |

#### Box 3.

Definitions for the Equilibrium Types in the SPS/ABD Games

| **Box 3. Definitions for the Equilibrium Types in the SPS/ABD Games** |  |
| --- | --- |
| **Nash equilibrium** | A *Nash equilibrium* (i.e., “no-regret” strategies) is a solution to a non-cooperative game involving two or more players, in which each player is supposed to be aware of the equilibrium strategies of the other players, and no player is better off by unilaterally changing own strategy. In other words, when each player has selected a strategy and no player may benefit from changing strategies whereas the other payers keep theirs fixed, then the current set of strategies and the consequent payoffs (fitness) constitute a Nash equilibrium. |
| **ESS (evolutionarily stable strategy)** | An *evolutionarily stable strategy* (ESS) is a strategy that, if adopted by a population of players in a given environment, cannot be invaded by any mutational (alternative) strategy, which is usually scarce initially. It is a Nash equilibrium that is “evolutionarily stable or resilient.” In other words, an ESS is a strong version of the Nash equilibrium, and the Nash equilibrium is a weak version of ESS. |
| **Pooling equilibrium** | A *pooling equilibrium* is an equilibrium in which, regardless of the types of sender with differing characteristic signals, the receiver shall choose the same response. In the case of the ABD game, Bob does not care whether or not Alice wears a mask when he makes his own decision. |
| **Separating equilibrium** | A *separating equilibrium* is opposite to pooling equilibrium: a receiver’s response can be different depending on the type of sender’s signal. In the case of ABD game, Alice’s signal influences Bob’s decision. |
| **Hybrid equilibrium** | A *hybrid equilibrium* is some combination of *separating* equilibrium and *pooling* equilibrium. |
| **Polymorphism** | A *polymorphism* is a mixed Nash equilibrium where sender mixes between being honest and dishonest signaling, and responder may also adopt a mixed strategy accordingly. Polymorphisms, which were ignored in biological signally until Huttegger & Zollman (2010) extensions of the original SPS game, correspond to hybrid equilibriums in this study. Polymorphic equilibrium allows for partially meaningful (honest) communication, and therefore grants the SPS much realism. |

#### Box 4.

Definitions for the Stability Properties in the SPS/ABD Games

| **Box 4. Definitions for the Stability Properties in the SPS/ABD Games** |  |
| --- | --- |
| **Lyapunov Stable** | Assume a dynamical system which is described with<br>$\dot{x} = f(x), \quad x \in R^n,$<br>there is an equilibrium solution $\bar{x} \in R^n$, such that $f(\bar{x}) = 0$. The equilibrium solution $\bar{x}$ is stable in the sense of Lyapunov stable, if for given $\varepsilon > 0$, there exists $\delta = \delta(\varepsilon) > 0$ such that, for any other solution $y(t)$,<br>$\ y(t_0) - \bar{x}\ < \delta \quad (t_0 \in R), \text{ then } \ y(t) - \bar{x}\ < \varepsilon \text{ for } t > t_0.$<br>Lyapunov stability is a rather weak requirement on equilibrium points. Especially, it does not require that trajectories starting close to the origin approach to the origin asymptotically. In a simplified interpretation, if the solutions that start out near an equilibrium point $x$ forever, then $x$ is Lyapunov stable. |
| **Asymptotically Stable** | A time-invariant system is asymptotically stable if all the eigen-values of its system matrix (**A**) possess negative real parts. In the case of asymptotic stability, there is a sphere $\mathcal{S}$, centered around $\delta = 0$ with the radius $r$, such that the response, once entered the sphere, converges to the origin. |
| **Neutrally Stable** | An equilibrium that is Lyapunov stable but not asymptotically stable is sometimes termed as *neutrally stable*. |

### Equilibriums and Stabilities of the ABD Game

Here we take advantage of the Huttegger & Zollman (2010) extensions and analytical works on the SPS game, and summarize the equilibriums of the ABD game in Table 2, and the payoff of the ABD game in Table 3, respectively.

To analyze the dynamic stabilities of ABD game, the *two-population* replicator dynamics model of SPS game can be formulated as follows, according to Huttegger & Zollman (2001):

Assuming *x*_*i*_ is the relative frequency of Alice’s strategy type *i*, and *y*_*j*_ is the relative frequency of Bob’s strategy type *j*, where *i, j*=1, 2, 3, 4, the replicator dynamics is as follows:

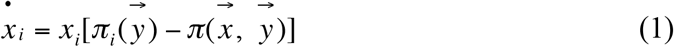

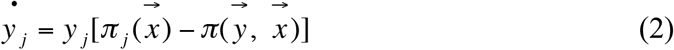

where 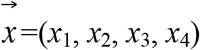 and 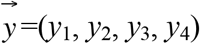, are the strategy sets of Alice and Bob respectively; 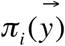 is the payoff of *i* strategy against 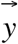 and 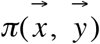 is the average payoff in Alice’s population; 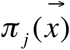 is the payoff of *j* strategy against 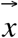 and 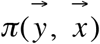 is the average payoff in Bob’s population.

We first interpret the equilibriums of the ABD game as follows:

### Strategy A is a Nash equilibrium

With the strategy pair A of (A_1_ *vs*. B_1_), Alice does not wear a mask only if she is healthy and Bob does not wear a mask only if Alice wears a mask. This strategy pair is a Nash equilibrium and is asymptotically stable (*see* Box 3 and Box 4 for the terminology interpretations). The rational for Alice may be that, if she was healthy, she would not need to wear a mask since there was no risk to spread virus, and Bob would wear a mask anyway to be cautious (in case Alice’s hid her healthy status). In this case, Alice is sending a message (“she is healthy”) by not wearing a mask, but Bob tends to be cautious; in particular, he does not necessary trust Alice’s signal.

Overall, the strategy pair ***A*** seems to mirror the behaviors of those who generally believe the benefits of mask wearing and tend to be cautious. Furthermore, since *A* is a Nash equilibrium, those believers do not regret for their decisions once they are committed.

### Strategy F is an ESS (evolutionary stable strategy) and is asymptotically stable

With the strategy pair F of (A_2_ *vs*. B_2_), Alice does not wear a mask only if she is infected, and Bob does not wear a mask only if Alice does not wear a mask. With this strategy, if Alice is infected, both Alice and Bob do not wear masks. This strategy is an ESS, which means it is resistant to ‘mutation’ or ‘invasion’—such as random change of individual behavior. However, it is not unbreakable. When external selection forces are too strong, the equilibrium may be overturned (Box 3 and Box 4). The apparent irrationality with this strategy is not totally out of touch with reality. For example, when Alice is overly concerned with possible social stigma associated with masking then she may choose not to wear a mask. Similarly, Bob may be concerned more with Alice’s perception on him than with the infection risk, particularly, when the overall infection probability is extremely low. Other possible scenarios in real world of such masking strategies may include the influences of misinformation or politics similar to those mentioned in Peeples (2020), and Bergstrom &West (2020). One may conceive that when the infection probability (*m*) and/or the strategic cost (*c*) (for nor honestly signaling) are dramatically raised, the equilibrium may be broken.

Overall, the strategy pair ***F*** seems to mirror the behaviors of mask skeptics. However, since ***F*** is an ESS, their belief may be changed when some of them “mutate” their behavior when the pandemic expands and ultimately they may be converted from skeptics to neutral or even believers.

### Strategy L is a pooling equilibrium

With the strategy pair ***L*** of (A_3_ *vs*. B_4_), Alice always wears a mask but Bob never wear a mask. The equilibrium is neutrally stable. Perhaps Bob felt protected if Alice wore one and it is unnecessary for him to wear one too, and mostly likely Bob could simply be a mask skeptic (given this is a pooling equilibrium).

Overall, the strategy pair ***L*** seems to mirror a divided world of mask believers and mask skeptics. The difference between the strategy ***L*** and previous described strategy ***F*** is that ***F*** is a world of mask skeptic (although they may be converted when the pandemic expands) and the ***L*** is a divided world of believers and skeptics (both populations may coexist). Again, such strategies may indeed exist in real world, similar to the scenarios described in Peeples (2020).

From a pure theoretic perspective, the equilibrium type ***L*** is more likely to become established because it has a larger basin of attraction, which measures the likelihood of evolving under standard evolutionary dynamics [Eqn. (1)-(2)].

### Mixed Strategies of J and K are pooling strategies and both are behaviorally equivalent

When Alice adopted strategy A_3_ (always masking), and Bob adopted a mixed strategy of B_2_ (not masking only if Alice does not mask) with probability (1−λ) and B_3_ (always masking) with probability λ, there is a pooling equilibrium that is neutrally stable. The condition for the equilibrium to occur is *d* > *k*[*ma* + (1 − *m*)*b*]. Bob’s mixed strategy is (1 − *λ*)*B*_2_ + *λB*_3_, and *λ* ≥ [1 − *c* /(*a* − *kd*)] determines the strategy of Bob. A scenario for achieving this equilibrium may be like this: Before dating, Bob was not aware of Alice’s healthy status. He either wore no mask when he spotted Alice did not wear a mask, or alternatively he always wore a mask (when he spotted Alice wore a mask, or he just took a fail-safe strategy).

Theoretically, when he hesitates, he can resort to λ to make an optimal decision, *i.e*., choose *B*_*2*_ when *λ* ≥ [1 − *c* /(*a* − *kd*)], *B*_*3*_ when *λ* < [1 − *c* /(*a* − *kd*)]. Since Alice always wears a mask, Bob would always wear a mask anyway, and the above rule for decision-making is moot in reality. Strategies ***J*** and ***K*** are behaviorally equivalent with each other.

Overall, the mixed strategies ***J & K*** seem to be closest to the universal voluntarily masking in real world. Some citizens might hesitate, but ultimately they followed the crowd.

### Mixed strategies of I and L are pooling strategies and both are behaviorally equivalent

When Alice adopted strategy A_3_ (always masking) and Bob adopted a mixed strategy of B_1_ (not masking only if Alice masks) with probability (*1*−*μ*) and B_4_ (never masking) with probability *μ*, where *μ* = 1 −[*c* /(*kd* − *b*)], there is a pooling equilibrium that is neutrally stable. Similar to previous J & K strategies, there is a pooling equilibrium that is neutrally stable, but here *μ* determines the strategy of Bob. Intuitively, since Alice always wears a mask, Bob would not wear a mask since he may believe it is unnecessary for him to wear one. Strategies ***I*** and ***L*** are behaviorally equivalent with each other.

Overall, the mixed strategies *I & L* appear to be closest to the voluntarily masking: people make rational decisions based on their own assessments of the risk they perceived.

### Polymorphic hybrid strategies

Alice adopted a mixed strategy of A_2_ (not masking only if infected) and A_4_ (never masking), and Bob also took a mixed strategy of B_2_ (not masking only if Alice does not mask) and B_3_ (always masking). With these options, there is a hybrid equilibrium that is *polymorphic*.

Overall, Alice seems to be a mask skeptic, and Bob appears to be variable. When he saw that Alice did not wear a mask, he chose to follow her to please her; but in other occasions, he ignored Alice’s message and chose to wear a mask. What’s interesting is that Bob might have a “split character” or bounded rationality: sometimes blindly following Alice, whereas being very cautious other times. This is obviously the most complex strategy interaction, and corresponding equilibriums heavily depend on the complex combinations of parameters as briefly discussed below. Here we first discuss how Alice and Bob make their decisions and then use simulations to further demonstrate some of the important properties of this strategy in the next section.

First, how would Alice make her decision? In fact, it is the parameter *μ* that determines her decision to choose either A_2_ or A_4_. Theoretically, when she hesitates, she can resort to *μ* to make an optimal decision, *i.e*., choose A_2_ when *μ* > *c* /(*b* − *kd*), A_4_ when *μ* < *c* /(*b* − *kd*). When *μ* = *c* /(*b* − *kd*), she tends to adopt mixed strategy *λA*_2_ + (1 − *λ*)*A*_4_.

For example, when *μ* > *c* /(*b* − *kd*), Alice would prefer to choose A_2_, she would *not* wear a mask (not worry of being infected) if she was already infected and wear a mask (worry of being infected) if she was not infected. This choice would suggest that Alice is a more selfish person. In contrast, when *μ* < *c* /(*b* − *kd*), Alice would never wear a mask anyway, perhaps worrying of being perceived as timid.

Second, how would Bob make his decision of B_2_ or B_3_ in response to Alice’s signal or lack of signal at all? Bob’s decision would depend on parameter λ. When *λ* > {*K*[*ma* + (1 − *m*)*b*] − *d*}/[(1 − *m*)(*kb* − *d*)], Bob would prefer to choose B_2,_ or B_3_ when *λ* < {*K*[*ma* + (1 − *m*)*b*] − *d*}/[(1 − *m*)(*kb* − *d*)]. When *λ* = {*K*[*ma* + (1 − *m*)*b*] − *d*}/[(1 − *m*)(*kb* − *d*)], he tends to adopt mixed strategy *μB*_2_ + (1 − *μ*)*B*_3_.

For example, when *λ* > {*K*[*ma* + (1 − *m*)*b*] − *d*}/[(1 − *m*)(*kb* − *d*)], Bob would prefer B_2_, he would not wear a mask if Alice did not either. In this case, Bob seemed to trust Alice’s signal. When *λ* < {*K*[*ma* + (1 − *m*)*b*] − *d*}/[(1 − *m*)(*kb* − *d*)], Bob would prefer B_3,_ ignoring possible prejudice against masking and always wearing a mask to avoid infection or being infected. This hybrid equilibrium is Lyapunov stable, which implies that the actual stability may vary depending on the values of parameters. In other words, the stability of equilibriums may be broken when conditions such as infection probability and/or cost change dramatically. Fig 2 shows an example of the decision-making in the phase space of the polymorphic hybrid equilibrium, obtained from the simulation study explained below. The polymorphic hybrid strategies, arguably, represent the most sophisticated options, which reflect the dynamic (evolutionary) nature of the masking-or-not decision-making.

**Fig 2.**
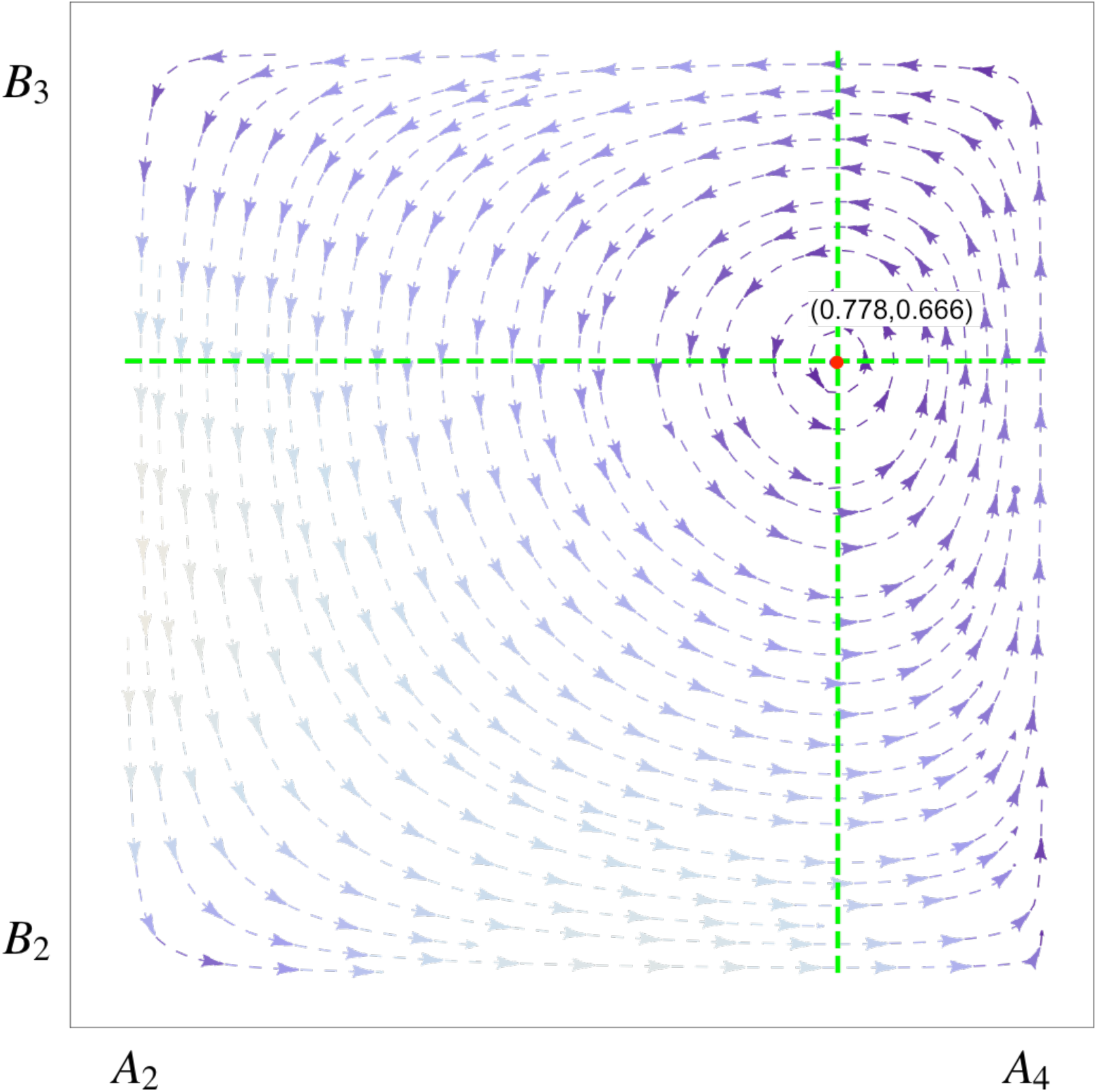
An example output from the simulation program, *i.e*., the phase portrait of the polymorphic hybrid equilibrium with the hybrid strategy of **Alice** (*A*_2_, *A*_4_) *vs*. **Bob** (*B*_2_, *B*_3_) under following parameter values: *a*=0.7, *b*=0.3, *c*=0.1, *d*=0.3, *k*=0.5 and *m*=0.4. Bob and Alice make their decisions based on (*λ, μ*)=(0.778, 0.666). If *μ*>0.666, Alice prefers to choose *A*_2_ over *A*_4_, as indicated by the direction of portrait arrow (to left). If *μ*<0.666, Alice prefers to choose *A*_4_ over *A*_2_, as indicated by the direction of portrait (to right). When *μ*=0.666, the mixed strategy of *A*_2_ and *A*_4_ is chosen. Similarly, Bob makes his strategic decisions based on the value of parameter *λ* >0.778, <0.778 or =0.778.

### Simulation Study

#### Simulation Procedures and Illustrative Graphs

The online supplementary information (OSI) include the codes in Python for simulating the replicator dynamics of the ABD game [Eqns. (1) and (2)]. Table S1-S6 in the OSI contain 6 MS-Excel sheets generated from the Python program. The sheets contain the ABD parameters and payoffs of the simulated equilibriums for each of the six equilibrium types. The lists of the equilibriums are obviously non-exhaustive, and were generated based on the necessary conditions (listed in Table 2) for those equilibriums. However, the full ranges of the parameters (all ranged between *0* and *1*) with step length=*0.1* were simulated, and therefore the simulated equilibriums can be considered as representative samples of the ABD equilibrium space.

Although the analytical solutions for the ABD game are available (Tables 2 & 3), they are far from intuitive, which is the main reason we performed above-described simulation experiments. Indeed, given their complexity, even with numerical simulations, it is still difficult obtain the full spectrum of complex behaviors transpired by the game model. The strength with simulations is that it can be helpful to get more intuitive interpretations for the behaviors. Below, we briefly discuss three simulated results, as displayed in Figs 2-5, as well as Table S7. All of the figures were drawn based on the simulation results in Table S1-S7.

Fig 2 shows an example for the dynamic properties of the equilibriums, *i.e*., the phase portrait of the polymorphic hybrid equilibrium with the hybrid strategy of Alice (A_2_, A_4_) *vs*. Bob (B_2_, B_3_) under following parameter values: *a*=*0.7, b*=0.3, *c*=0.1, *d*=0.3, *k*=0.5, and *m*=*0.4*. Bob and Alice make their decisions based on (*λ, μ*)=(0.778, 0.666), which is an equilibrium point as displayed as red spot in Fig 2. If *μ>0.666*, Alice prefers to choose *A*_*2*_ over *A*_*4*_, as indicated by the direction of portrait arrow (to left). If *μ<0.666*, Alice prefers to choose *A*_*4*_ over *A*_*2*_, as indicated by the direction of portrait (to right). When *μ =0.666*, the *mixed strategy of A*_*2*_ and *A*_*4*_ is chosen. Similarly, Bob makes his strategic decisions based on the value of parameter *λ >0.778, <0.778 or =0.778*. Similar phase portraits for other types of equilibriums could be drawn based on the results in Table S1-S6.

Fig 3 exhibits an example demonstrating the influences of the infection probability (*m*) and strategic cost (*c*) of signaling on Alice’s or Bob’s payoff (fitness) in the case of polymorphic hybrid equilibriums. Higher infections and costs correspond to lower payoffs, and vice versa, lower infections and costs lead to higher payoff, which should be expected. The effects of other parameters such as relatedness (*k*) on the payoffs could also be plotted based on the simulated results in Table S1-S6.

**Fig 3.**
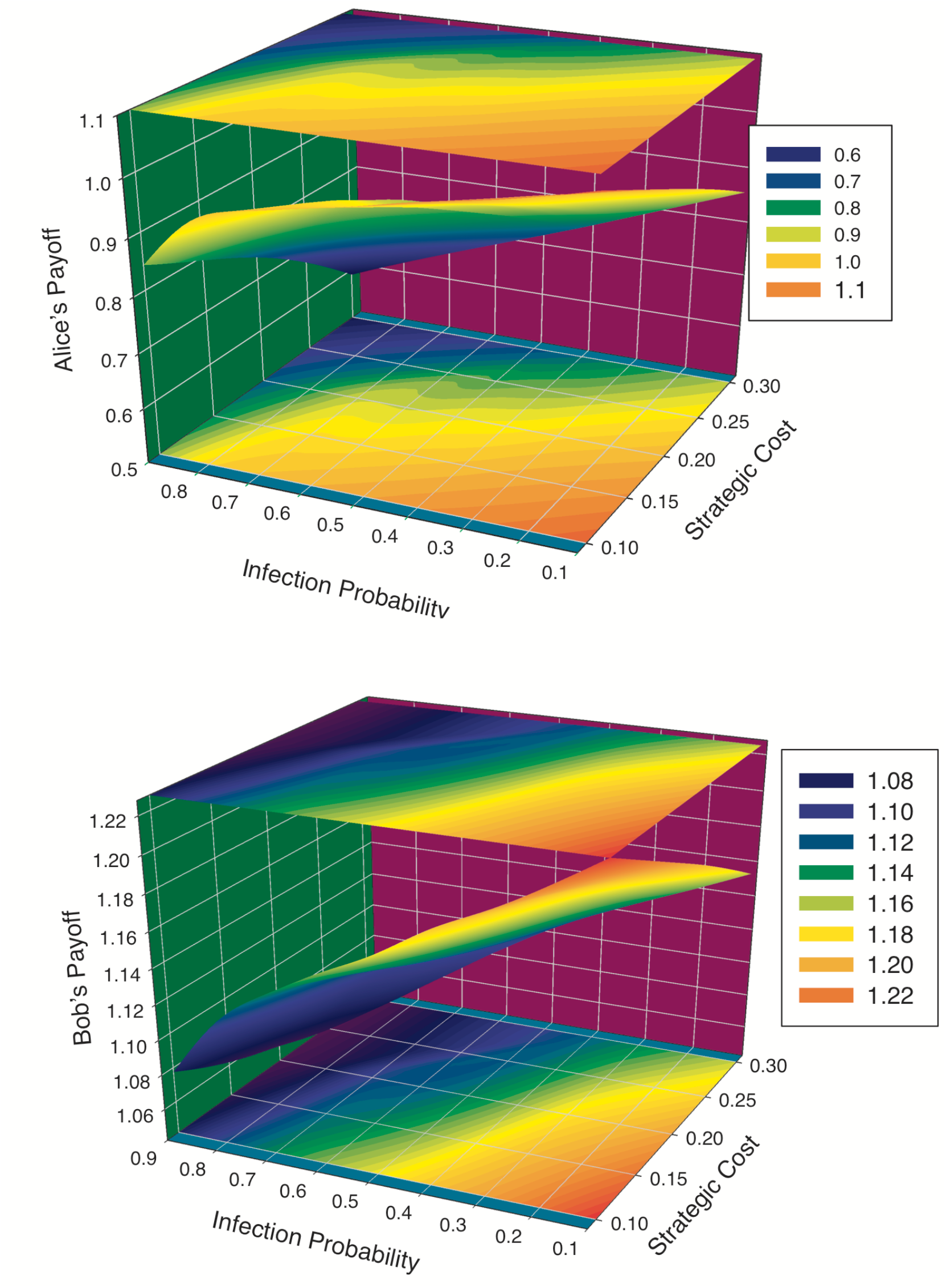
Simulations showing the influences of infection probability and strategic cost on the players’ payoffs (top for Alice’s payoff and bottom for Bob’s payoff): When the cost and infection are high, Alice’s payoff approximates to the floor, and vice versa Alice’s Payoff approximates to the ceiling. Bob’s payoff exhibited similar trend. Indeed, their payoffs are positively correlated as further illustrated in Figs 4 & 5.

Fig 4 displays the relationships between Alice’s payoff and Bob’s payoff. We fitted both linear and quadratic (parabolic) regression models to the payoff data in Table S1-S6, and the fitted model parameters were listed in Table S7 for all six equilibrium types listed in Table 2. Overall, the relationships follow the parabolic (quadratic) model for four of the six equilibrium types, including A, F, J-K and hybrid, follow the simple linear model for the remaining two equilibrium types (*i.e*., L, I-L). It seems that the linear relationship between Alice’s and Bob’s payoffs could be attributed to Alice’s straightforward “always masking” strategy, which might “smooth” their payoff relationship. Indeed, in the case of the two linear models, most data points straightly fall on the fitted straight lines, where the data points with the parabolic curve are more scattered and suggesting stronger fluctuations between Alice’s and Bob’s payoffs. The fluctuation is obvious in Fig 5 as explained below.

**Fig 4.**
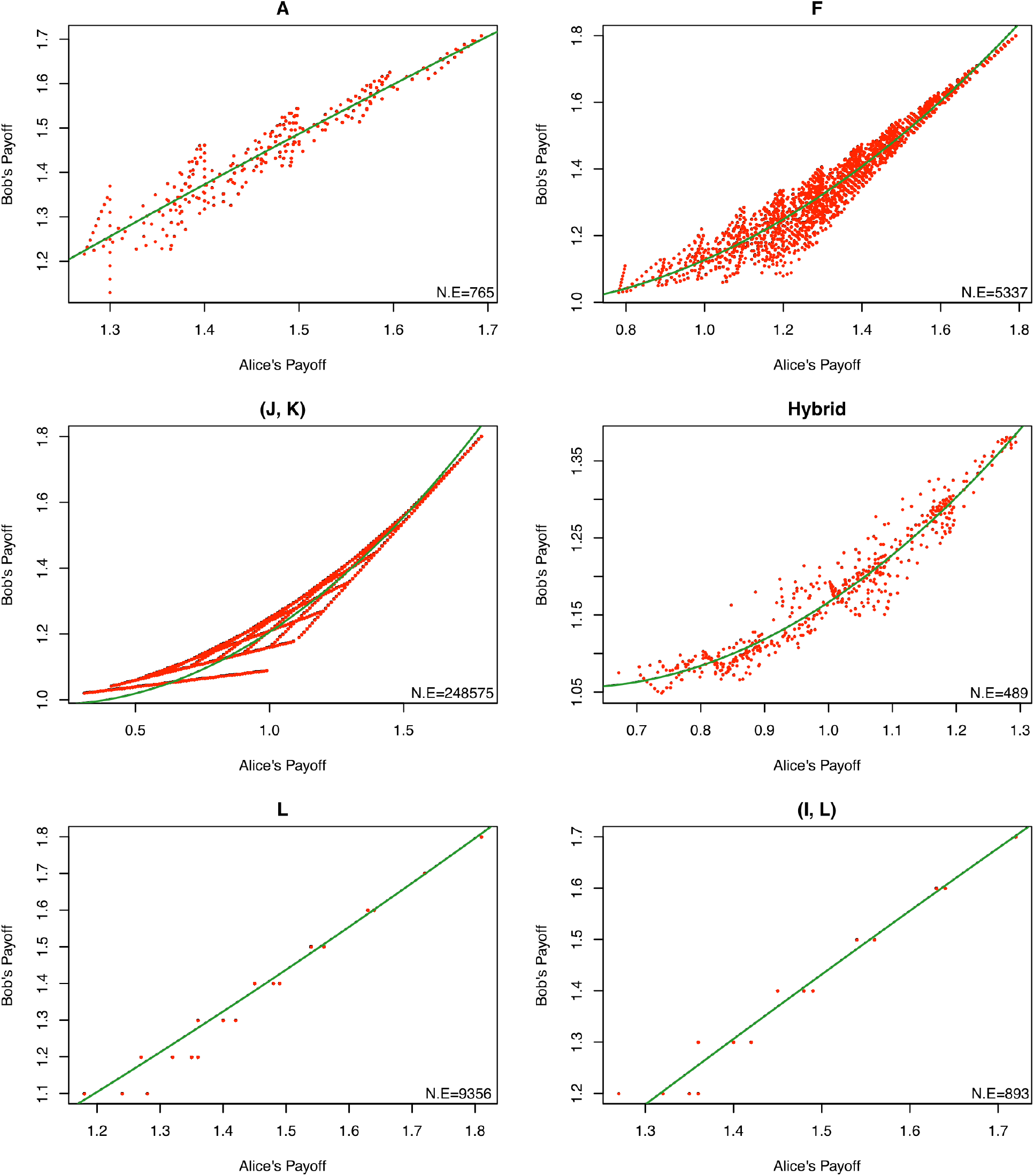
Simulations illustrating the relationships between Alice’s and Bob’s payoffs: parabolic relationships with equilibriums (A, F, JK, and hybrid: the top four), and linear relationships with equilibriums (L & IL: the bottom two). Notice that the N.E at the right corner of each graph shows the *number of equilibriums* obtained from the simulations and was used to draw the graph, and two linear graphs appear to have fewer data points due to points overlaps. We postulate that the two simple linear relationships may be attributed to Alice’s straightforward strategy (“always masking”).

**Fig 5.**
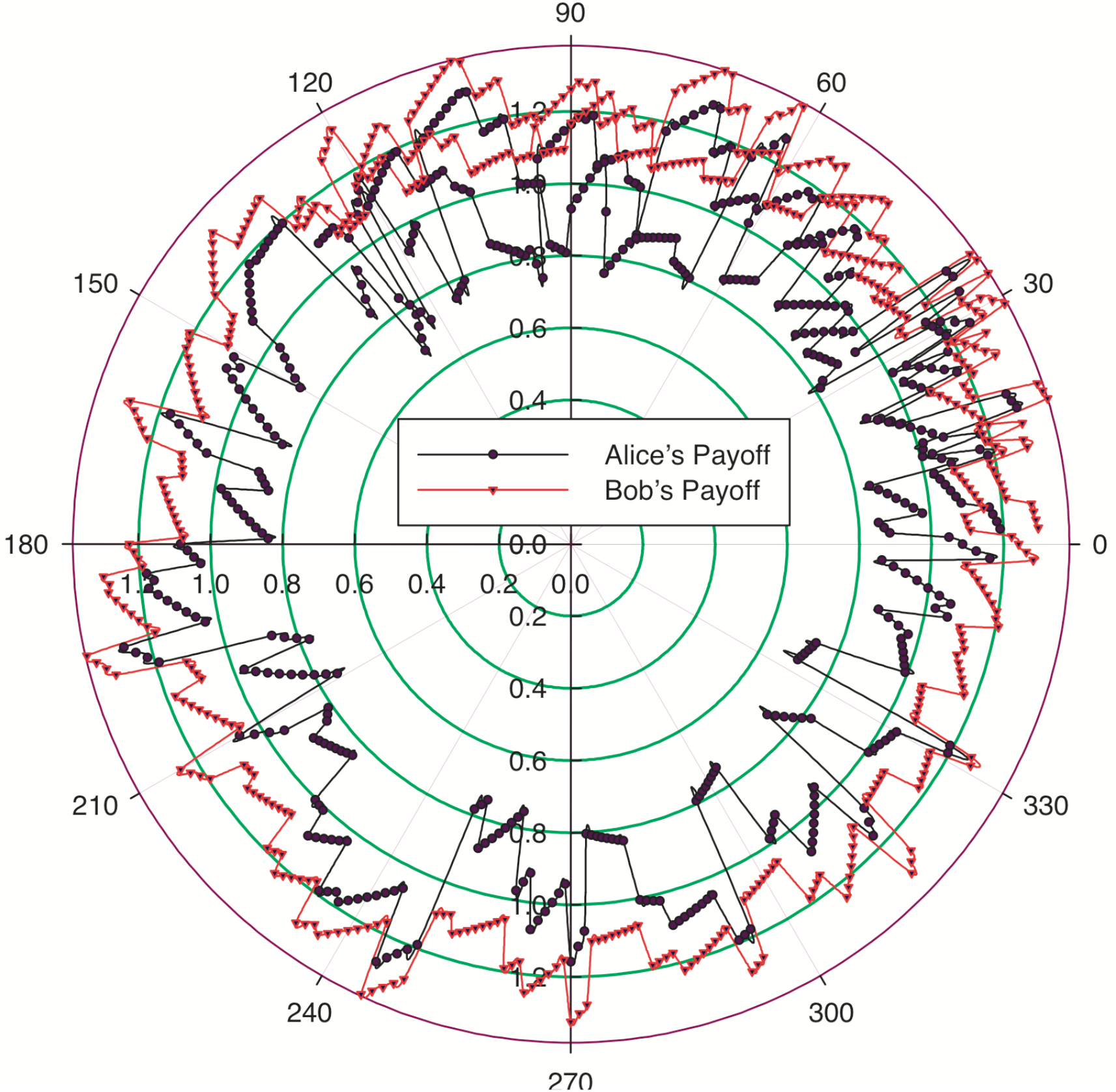
A polar coordinate graph showing the correlation between Alice’s and Bob’s payoffs with the polymorphic hybrid strategies: the radial coordinate showing the payoffs and the angular simply showing the sequencing numbers of each pair of payoffs, which does not actually have particular order (could be random) and is not really interesting. What is interesting is, we observed, that the relationship on a coarse scale (macroscopic) may be parabolic or even linear (*see* Fig 4), but the relationship on a fine scale (microscopic) may be complex nonlinear fluctuations, suggesting subtle intricacies between the payoffs and mirroring the complex idiosyncratic behaviors surrounding the ABD game.

In Fig 5, the relationship between Alice’s and Bob’s payoffs for the polymorphic hybrid equilibrium was redrawn with polar coordinate system. The polar coordinate more conspicuously illustrated the locally fluctuating payoff relationship, rather than globally stable parabolic relationship as shown in Fig 4. In other words, the relationship on a coarse scale (macroscopic) can be parabolic or even linear (*see* Fig 4), but the relationship on a fine scale (microscopic) seem to be complex nonlinear fluctuations, suggesting subtle intricacies between Alice’s and Bob’s payoffs and mirroring the complex idiosyncratic behaviors surrounding the ABD game.

#### Social efficiency deficit (SED) approach to the ABD Game Strategies

During the peer-review process, both anonymous reviewers have suggested additional demonstrations of the ABD-game behaviors based on the social efficiency deficit (SED) approach (Arefin *et al* 2020; Tanimoto 2015, 2021; Wang *et al*. 2015). The SED was defined as the difference between a play’s (maximal) payoff (fitness) achieved in a social optimum and an evolutionary equilibrium (averaged from neighboring equilibrium points) (Arefin *et al*. 2020, Tanimoto 2015, 2021, Wang *et al*. 2015). The concept was borrowed from the studies of social dilemma. The greater the SED is, the greater is the potential for the betterment of society while transitioning from the evolutionary equilibrium to the social optimum. In other words, a larger SED value implies smaller level of social dilemma or larger cooperative tendency (smaller rebellious tendency). In contrast, a smaller SED value implies larger level of social dilemma or smaller cooperative tendency (larger rebellious tendency) (Arefin *et al*. 2020, Tanimoto 2015, 2021, Wang *et al*. 2015). We suggest using the relative SED, which can be defined as the original SED divided by the social optimum value (SOV). Obviously, the relative SED can be directly used to compare various equilibrium strategies.

To apply the approach, we randomly picked one set of ABD (Alice and Bob’s dilemma) parameters for each of the equilibrium types, and then compared them from the perspective of social efficiency deficit (SED). From our simulations of the five equilibrium types (Table S1-S6), we randomly selected five sets of ABD parameters, and computed their SED values (Table 4). It was found that most SED values are relatively small, suggesting that the level of social dilemma or rebellious tendency is rather high. This reflects the reality of split society on the masking behavior. The hybrid strategy has the highest SED, suggesting that the strategy is least controversial, which is consistent with the mixed nature of the strategies—compromising between players. It should be mentioned that the SED approach demonstrated with the parameter values exhibited in Table 4 can be applied to other simulated parameter values presented in online supplementary Tables S1-S6. Overall, the SED approach can act as an effective supplement to our ABD game model for obtaining further insights on the masking behaviors corresponding to various equilibriums.

**Table 4.** Demonstrating the computation of the social efficiency deficit (SED) for the various equilibrium types with selected parameter (*a, b, c, d, k, m*) values

| Equilibrium Type | Parameters ($a, b, c, d, k, m$) | Social Optimum Values (SOV) | Evolutionary Equilibrium Values | Social Efficiency Deficit (SED) | SED/SOV |
| --- | --- | --- | --- | --- | --- |
| $A$ | $a=0.5, b=0.1, c=0.1, d=0.3, k=0.9, m=0.2$ | 3.4960 | 3.4453 | 0.0507 | 0.015 |
| $F$ | $a=0.3, b=c=d=0.1, k=0.9, m=0.3$ | 3.6100 | 3.5245 | 0.0855 | 0.024 |
| $I$ and $L$ | $a=0.3, b=c=0.1, d=0.2, k=0.9, m=0.7$ | 3.4200 | 3.3440 | 0.0760 | 0.022 |
| $J$ and $K$ | $a=0.2, b=c=d=m=0.1, k=0.9$ | 3.6100 | 3.5910 | 0.0190 | 0.005 |
| Hybrid | $a=0.7, b=0.3, c=0.1, d=0.2, k=0.3, m=0.7$ | 2.3400 | 1.8460 | 0.4940 | 0.211 |

#### Basin of Attraction of the ABD Equilibriums

A dynamical system such as the ABD evolutionary game may possess the so-termed *attractors*, which are portions or subsets of the phase space (*e.g*., the space illustrated in Fig 2). An attractor’s *basin of attraction* measures the size of the region in the phase space, over which the system states will asymptotically be iterated (evolved) into the attractor, regardless of the initial condition. Further interpretation on the basin of attraction is referred to Huttegger & Zollman (2010). We chose to estimate the basin of attraction for the polymorphic hybrid equilibrium of the ABD game. Table 5 exhibited one set of the basin estimates for a randomly selected set of values of the ABD parameters.

**Table 5.** A demonstrative example illustrating the influence of strategic cost (*c*) on the evolution of polymorphic hybrid equilibrium based on estimation of basin of attraction: When *0*<*c*<*0.187*, {the mixed strategies: (A2, A4); (B2, B3)} can achieve the hybrid equilibrium.

| Alice and Bob Strategy* | Proportion of Mixed Strategies** | Payoff | Selection_Force<br>=(Payoff-Mixed_Payoff) | Mixed_Payoff***<br>=Weighted_Payoff |
| --- | --- | --- | --- | --- |
| A1: Not masking only if healthy | 0.01 | $0.7918-0.3c$ | $-0.2845+0.3880c$ | $1.0762-0.6880c$ |
| A2: Not masking only if infected | 0.95 | $1.0850-0.7c$ | $0.0088-0.0120c$ | |
| A3: Always masking | 0.02 | 0.7356 | $-0.3406+0.6880c$ | |
| A4: Never masking | 0.02 | $1.1412-c$ | $0.0650-0.3120c$ | |
| B1: Not masking only if Alice masks | 0.02 | $1.0942-0.206c$ | -0.0355 | $1.1297-0.2064c$ |
| B2: Not masking only if Alice does not mask | 0.80 | $1.1318-0.206c$ | 0.0021 | |
| B3: Always masking | 0.17 | $1.1260-0.206c$ | -0.0037 | |
| B4: Never masking | 0.01 | $1.1-0.206c$ | -0.0297 | |

\* The values of parameters (other than $c$ ) used are: $a=0.7$ , $b=0.3$ , $d=0.2$ , $k=0.3$ , $m=0.7$ , $\lambda=0.98$ , $\mu=0.83$ , which were selected from Table S6. Note that the payoff (Column #3) is negatively correlated with $c$ , which suggests that a larger strategic cost ( $c$ ) may discourage the evolution of the hybrid strategy equilibrium.
\*\* The proportion was set randomly with a constraint of the total proportion equal to unit (1 or 100%) for each player. For example, Alice takes a mixed strategy of A1, A2, A3, and A4 with probability of 0.01, 0.95, 0.02, and 0.02 respectively, and the total is equal to 1. In terms of evolutionary game, Alice and Bob represent two respective populations with mixed strategies as specified by the *Proportion* (Column #2) in Table 5. As an individual, Alice (Bob) takes mixed strategy with probability specified by the probability (=Proportion).
\*\*\* *Mixed\_Payoff=Weighted\_Payoff* weighted by the *Proportion*, i.e., the payoff of each strategy is weighted by the strategy's proportion and then summed up.

The selection force, which is defined as the difference between a strategy’s *Payoff* and the *Mixed_payoff* (from the mixture of all his or her strategies), represent the tendency (driving force) for a player to select a particular strategy, which is a simple linear function of strategic cost (*c*) (when other parameters are fixed) as displayed in Table 5.

From Table 5, one can infer the following insights: When 0<*c*<0.187, the payoff that Alice should select A2 (Not masking only if infected with payoff>0.9541) is slightly lower than selecting A4 (Never masking with *payoff*>0.9542), but significantly higher than selecting the other two strategies (A1 or A3). This means that when the cost of infection is low (*c*<0.187), Alice would tend to select behavior of “*not masking*” even if the infection risk is high (*m*=0.7 in this case). Second, the payoff that Bob should select B2 (Not masking only if Alice does not mask with *payoff*>1.092) is the highest, and his next favorable strategy is B3 (Always masking with *payoff*>1.0837), but their *payoff* difference is insignificant. This means that when the strategic cost (*c*) of infection is low, Bob would still select masking.

In real world, the above interpretations of Alice and Bob’s behaviors appear counter-intuitive. However, one possible explanation for their apparently contradictory behaviors could be related to the politics of COVID-pandemic, for example, the ‘left’ (always masking, like Bob in this case) *vs*. ‘right’ (never masking, like Alice). Nevertheless, beyond the pandemic politics, as humans, we have certain level of cooperative tendency. For Alice and Bob, their romantic relationship (which is represented by relatedness parameter *k*) should also play a role in their decision-making.

## Conclusions, Discussion and Perspective

### Conclusions and Discussion

It has been argued that one of the most important reasons why we humans have come to dominate the earth is attributed to our exceptional evolutionary capacity for decision-making (Samson 2020). Although on macroscopic scale, we have been enormously successful from selecting the right food and shelter, to devising complex economic strategies and effective public health policies, it is fair to say, we occasionally make expensive and painful mistakes on both an individual and a group level (Samson 2020). The COVID-19 pandemic has been one of the biggest challenges to the health and well-beings of humans in recent decades. It is obviously in the best interests of whole society and each individual to make right decisions to minimize the pandemic impacts. The challenging decision-making in facing the COVID-19 pandemic crisis involves both groups and individuals, and history tells us groups are not necessarily more likely to make right decisions, especially when the *communication* and *interactions* between individuals could not be controlled (Sunstein & Hastie 2015, Samson 2020). The bounded rationality and lack of full honesty may be inherent with *Homo sapiens*, which makes some apparently simple decision-making such as masking disproportionally complex. The challenge seems particularly real in the era of disinformation (*e.g*., Bergstrom & West 2020). It was these properties that prompted us to apply the SPS game and underlying handicap principle, which has achieved enormous success in analyzing the evolution of animal communications, to study the ABD problem. Since we are only interested in strategic decision-making on masking, it is natural for us to focus on the stable equilibriums and corresponding payoffs under variously possible strategies. Tactical or operational level analyses are obviously beyond the scope of this article.

In previous sections, we formulated the ABD as an evolutionary game, specifically as a SPS game. There are 16 possible options or strategy combinations given that SPS game is an asymmetric four-by-four strategic-form game. We summarized a total of six types of equilibriums (Table 2) from the 16 possible strategic combinations and their payoffs (inclusive fitness) (Table 3). Those equilibriums can be distinguished as separating (signaling), pooling, and polymorphic hybrid equilibriums, and they may have different behavioral and payoff properties (*see* Box 3, Box 4 and Table 3). Furthermore, their stabilities can be dramatically different (Table 2), which may have different theoretical and practical ramifications. Theoretically, those ramifications have been extensively studied and well documented in the existing literatures (*e.g*., Maynard-Smith & Harper 2003, Huttegger & Zollman 2001, Bergstrom & Lachmann 1997, 1998, Biernaskie et al 2018, Madgwick & Wolf 2020, Whitmeyer 2020). However, their practical ramifications are much more complex, which we briefly discuss below.

In our opinion, the ABD provides a powerful abstraction for analyzing the masking behavior in the COVID-19 pandemic. However, the ABD can only reveal possible behavior types (game strategies). To fully understand various masking (or not masking) behavior decisions, we must also consider the cultural and humanity dimensions of the problem. While medical science studies have generally support the masking or even universal masking strategies (*e.g*., Klompas *et al*. 2020, Gandhi *et al*. 2020, Peeples 2020), in reality, from the very beginning of COVID-19 pandemic until today, protests have never stopped in some regions of the world. Indeed, masking still seems to be one of few most controversial issues in public health policy surrounding COVID-19 (Peeples 2020). In the remainder of this article, we try to shed light on the ABD of masking from the perspective of behavior economics.

According to behavior economics (Samson 2014, 2020), it is not always true than humans are self-interested, benefits maximizing, and costs minimizing with stable preferences. Our minds may only possess *bounded rationality*, which suggests that human rationality is limited by brain’s information processing capability, insufficient knowledge feedback, and time constraint. For example, our thinking and decisions may be strongly influenced by readily available information in memory, automatically produced affection, and salient information in the environment. Humans live in the moment, often resist to changing and may be bad in predicting future behavior. Last but not the least important, humans are social animals with social preferences, including those embedded in trust, reciprocity and fairness. We are also emotional animals and are subject to social norms, caring reputation and self-consistency (Ma 2015a, 2015b, Samson 2014, 2020).

An obviously different attitude has emerged between the Eastern and Western towards the using of facial mask at the very beginning of the COVID outbreaks. We postulate that the difference might be related to holistic *verse* analytic thinking styles. It has been argued that the differences in thinking styles may have a profound influence on the tensions between the psychology of *Homo economicus* and *Homo sapiens*, being relatively weaker in Eastern-Asian cultures (Nisbett *et al*. 2001). The holistic reasoning is more likely context-dependent, in which people tend to use their intuition more if it is in conflict with formal rationality and tend to accept variations across scenarios (Nisbett *et al*. 2001, Samson 2014). This may explains relatively less resistance to the advocacy of using masks in public places in the Eastern Asian.

It might be disappointing that we can produce few concrete recommendations to the general public from the present study regarding the masking dilemma, other than highlighting its complexity and its multidimensional nature (including science, culture, information availability, and *etc*), for which existing studies have presented confirmative recommendations in general (*e.g*., Peeples 2020). Instead, our contribution, hopefully being intellectual, lies in the formal abstraction of the problem and presented formal strategic options as well as their stabilities and payoffs in 16 possible strategic options (scenarios) and 6 types of equilibriums. Whereas the number of scenarios (16), equilibrium types (6) with various stability features may vary with other alternative abstract models, we believe our ABD game model is sufficiently general for further scientific investigation of the dilemma from a multi-dimensional perspective. As to the reason why we cannot generate tangible recommendations at this stage, it becomes clear if one notices how the society has been dealing with the impact of cigarette smoking, even after the hazards of second-hand smoking have been revealed by many scientific studies (Samson 2014, 2020). If we choose to make one recommendation, our suggestion would be that *nudging* could be a more effective strategy in advocating masking. In recent years, nudging or nudge philosophy has been used in many public-policy devising such as consumer welfare (Thaler & Sunstein 2008). The nudge philosophy treat people as *Homo behaviouralis*, shifted from *Homo economicus*, in which policy or advocacy is designed to modify the context in which decisions are made without changing the constraints faced while the freedom of choice is preserved (Samson 2020). For example, rather than posting “no mask, no service”, perhaps an alternative post with “please keep 2 meters of distance if you forget to bring mask” could be equally effective.

In conclusion, despite the demonstrated complexities from our ABD game model, it seems that the six types of equilibriums theoretically derived from the game model adequately explain the diverse “norms of masking” evolved in much of the world with the progression of COVID-19 pandemic. Those norms represent diverse behaviors of mask believers, skeptics, converted, universal masking, voluntarily masking, coexisted and/or divided world of believers and skeptics, and their evolutions (formations) have obviously influenced by various scientific, social and other complex factors, and most importantly the severity of the pandemic. They are of different level of stabilities (resilience) at present and they are expected to continue evolve with the progression of the pandemic. As a science paper, we stick to the validity of mathematic logic underlying the ABD game model. Nevertheless, we do not pronounce a position for the right or wrong of those norms (including those that may be counterintuitive, unnatural or irrational ones) since we believe mutual respects and accommodations are part of the humanity. We are hoping that the breakthroughs in medical sciences such as vaccines or new treatments will ultimately make the issue of masking moot. The only regret (in our opinion) might be that Alice and Bob are likely to stick to working at home even after their dilemmas disappear! Of course, we wish them a lifetime of love and happiness!

### Perspective

It is the summer of 2021 already when we are revising this manuscript, while the COVID-19 pandemic has been going on wave after wave for near two years, despite the expanding vaccination efforts worldwide. The emergence and spreading of significantly more vicious *δ*-variant and somewhat disappointing protective efficacy of vaccines have prompted some countries (regions) to reenact some level of mask mandates, together with vaccination promotions and even mandates, while uncertainties surrounding the pandemic refuse to fade away. Public attitudes to masks have gone through about-face changes in many parts of the world, and mostly turned to accepting it as an effective personal protection equipment. From the perspective of ABD game modeling, the rising infection risk (*m*) should have shaped the evolution of masking behaviors since the start of the pandemic; yet we believe other social, cultural, economic and political factors have certainly played significant roles too.

Through this article, we have minimized the discussion on the politics surrounding masking behaviors/policies and COVID-19 at large. Nevertheless, in reality, politics are hardly avoidable in making public health policies. We realize that some of the peculiar, idiosyncratic behaviors simulated by the ABD game might be rightly explained by science denialism, politics or political economy of pandemics. In fact, during the submission processes of this manuscript, we were advised by an anonymous expert reviewer in another journal to formulate the ABD game as “one in which the signaler’s state is instead her political leaning (left or right, say), …” We do believe that was an excellent idea and may pursue it in future.

While avoiding or minimizing politics in the science of anti-pandemic is crucial for making sound public health policies such as masking, vaccination, quarantines, and/or lockdown, it is equally important to avoid the “counter-revolution of science” (Hayek 1955). The “counter-revolution” of science refers to the attempt to remove the human factor in order to obtain objective, strictly controlled results by using the so-termed hard science approaches, or the attempt to measure human action itself with the soft science approaches. The limitations of such attempts can be particularly serious when the scientific reasoning is based on incomplete information or knowledge. It is for this reason, we caution that the results of ABD modeling such as the equilibriums should only be used as explanatory/exploratory purposes at strategic level, rather than used for advising public at tactic levels.

We argue that the methodology we used to model masking behaviors with the ABD game should also be inspirational for exploring other public health policies (measures) such as social distance, lockdown/reopen and quarantines (abbreviated as DLQ hereafter). These measures are designed to physically contain the spreading of pathogens by isolation (distancing or containment) and can be characterized as either “open *vs*. closed”, *i.e*., without *vs*. with enforcing isolation (containment) measures such as DLQ. Obviously, masking also belongs to isolation. According to the metapopulation (*i.e*., *population* of local populations) theory (Citron *et al*. 2021, Ma 2020), infectious diseases such as COVID-19 can be modeled as a metapopulation of infectious pathogens, *i.e*., consisting of many local (regional) populations of pathogens (carried by human hosts) such as the local or regional outbreaks of COVID-19 (*e.g*., outbreaks in different countries). Also according to classic ecological theories (Hilker *et al*. 2009, Friedman *et al*. 2012, Ma 2020), the extinctions of local populations can be common events, although the global metapopulation is usually stable and resilient against global (total) extinction. Hilker *et al*. (2009) demonstrated theoretically that the disease dynamics could be rather sensitive to perturbations such as disease control methods even when the basic reproductive number (*R*_0_) exceeds *1* significantly (*R*_0_>>1). *R*_0_ is often used to evaluate the potential for disease invasion and persistence, to forecast the extent of an epidemic, and to infer the impact of interventions and of relaxing control measures (Shaw & Kennedy 2021).

The Allee effect is named after animal ecologist W. C Allee, and it refers to a theoretic threshold of population size, above which population growth may accelerate and below which population may go extinct (Kramer *et al*. 2017). Friedman *et al*. (2012) investigated the interaction (relationship) between the Allee effect of disease pathogens and the Allee effect of hosts (*e.g*., humans); their theoretic analysis was aimed to answer a question of fundamental importance in epidemiology: Can a small number of infected individual hosts with a fatal disease drive the host population to go extinct (assuming the host population is subject to the Allee effect)? Ma (2020) estimated the fundamental migration (dispersal) number (migration rate) of COVID-19 and different countries (regions) may have different numbers. He also estimated the population aggregation critical density, the threshold for aggregated infections to occur (Ma 2020). These studies suggest that there could be thresholds (tipping points) at which local pathogen populations may go extinct, depending on the dynamics of host-pathogen system, impacted by disease control measures such as the previous defined open/closed polices. Of course, it should be a goal of public health policies to possibly drive the pathogen populations (such as SARS-COV-2), at least locally, to go extinct, while keep the host population absolutely safe from the risk associated with the Allee effect of the hosts. As demonstrated by Citron *et al*. (2021) with their metapopulation dynamics modeling, the human movement has a critical impact on the global spreading of infectious diseases across large geographical distances. Therefore, containment or distancing measures that can regulate human movement behaviors should be able to play a critical role in containing the pandemics such as COVID-19.

Now imagine that there are two neighboring countries *A* and *B*, who may adopt different anti-pandemic strategies, *open* (without adopting DLQ measures) or *closed* (with DLP measures). They may or not cooperate with each other by adopting the same or different strategies. Similar to previously discussed ABD game for masking behaviors, their tendency to cooperate can be measured by parameter *k*, indicating their shared interests. The strategic cost (*c*) in the ABD game can be used to characterize the consequence (such as the fatality and/or economic loss) of failure in controlling local outbreaks. The parameter (*m*) can be treated as a probability function of *R*_0_ (basic reproductive number) of the pathogen. Parameters *a, b*, and *d* could be treated as probability functions that may lower the resilience (survivability) of society. With such an evolutionary game model inspired by the ABD game (let us call it “open or close dilemma”: OCD), we can expect that the complex behaviors of two countries similar to Alice and Bob be generated from the OCD model. Both countries can, in fact, represent two “metapopulations” of countries (local populations), and within each metapopulation, different countries (local populations) may actually adopt different strategies (similar to Alice and Bob each has four different strategies). With the above conceptualization, the suggested OCD model can be used to analyze the interactions of two countries (metapopulations); and the effects of the key epidemiological and ecological parameters such as the thresholds of Allee effects and *R*_0_ can be investigated at regional/global scales.

Within the context of the suggested OCD game scheme, many countries may adopt coexistence (between humans and the pathogen) strategy, some may pursue zero-infection goal dynamically, and still others may adopt *laissez-faire* strategy (*e.g*., passive herd immunity). Intuitively, zero-infection strategy is apparently unrealistic, but it should be possible locally at the minimum, if Allee effects of the pathogen can be exploited effectively. In fact, the eradication of smallpox is a successful example of zero infection strategy. Each of the strategies may possess some unique merits, and some of which could be exclusive. There may not be a simple criterion (or even a set of criteria) to evaluate various strategies objectively given the enormous complexities of the scientific and technological, geographic, cultural, economical and political factors underlying the host-pathogen dynamic system. For example, Sy *et al* (2021) estimated the *R*_0_ of 1,151 US counties with the medium of *R*_0_=1.66, *ranged* from 0.38 to 12.44. One particularly important point we would like to emphasize from their findings is that the *range* of [0.38, 12.44] was largely dependent on local (county) population densities Their *R*_0_ numbers were estimated before more contagious SARS-CoV-2 variants emerged. With the recent *δ*-variant, the upper limit of the range might be one order of magnitude higher, *i.e*., possibly >100 in some of the counties with highest population densities in the US, according to some recent reports on the *R*_0_ of *δ*-variant. Of countries, there are many countries in the world that have higher population densities than the US. For example, while the inter-residence distances in rural USA are usually hundred of meters away, and the distance can be as few as a few meters in the apartment settings in super big cities or in countries with higher population densities. The distance differences could easily exceed 2-3 orders of magnitude. If this assumption of the difference in residence distances holds, and if the “amplification” effects of population density (especially the residence settings) discovered by Se *et al*. (2021) is also applicable outside the US, then the *R*_0_ of SARS-CoV-2 (including *δ*-variant) in some densely populated countries may be raised by another 1-2 orders of magnitudes. The higher *R*_0_ also implies that the occurrences of super-spreading can be more frequent and severer.

Given potentially 2-3 orders of differences in *R*_0,_ the same or similar co-existence (tolerance) policy in different countries can produce dramatically different consequences. While co-existence policy in a sparsely populated country may be a rational and even advantageous policy, the same policy may lead to disastrous outcomes in a densely populated country. Obviously, population density is certainly not the only major factor that influences *R*_0_, and of course *R*_0_ is not the only critical parameter of pandemics. For example, the threshold of Allee effects is another critical parameter for suppressing pandemic or eradicating a pathogen. Therefore, exploring the policies and strategies for fighting pandemics should sufficiently consider complex scientific, technological, and socioeconomic factors that shape the pathogen-host dynamic systems. Game-theoretic models such as ABD game can offer important cognitive tools for the decision-making in fighting pandemics.

## Data Availability

This is a theoretical analysis, and only involves simulated data.

## Acknowledgements

This study received funding from the National Science Foundation of China (Grant #31970116); the Cloud-Ridge Industry Technology Leader Grant. We appreciate the assistance from HJ (Daisy) Chen and ST Li from Computational Biology & Medical Ecology Lab, Chinese Academy of Sciences in preparing the illustrative figures and checking the tables.

